# Disentangling the roles of human mobility and deprivation on the transmission dynamics of COVID-19 using a spatially explicit simulation model

**DOI:** 10.1101/2020.11.25.20144139

**Authors:** C.J. Banks, E. Colman, T. Doherty, O. Tearne, M. Arnold, K. Atkins, D. Balaz, G. Beaunée, P. Bessell, J. Enright, A. Kleczkowski, G. Rossi, A.-S. Ruget, R.R. Kao

**Affiliations:** Roslin Institute, University of Edinburgh; The Animal and Plant Health Agency, Weybridge, Surrey; Usher Institute, University of Edinburgh; INRAE, Nantes, France; School of Computing Science, University of Glasgow; Mathematics & Statistics, University of Strathclyde; Royal (Dick) School of Veterinary Studies

## Abstract

Restrictions on mobility are a key component of infectious disease controls, preventing the spread of infections to as yet unexposed areas, or to regions which have previously eliminated outbreaks. However, even under the most severe restrictions, some travel must inevitably continue, at the very minimum to retain essential services. For COVID-19, most countries imposed severe restrictions on travel at least as soon as it was clear that containment of local outbreaks would not be possible. Such restrictions are known to have had a substantial impact on the economy and other aspects of human health, and so quantifying the impact of such restrictions is an essential part of evaluating the necessity for future implementation of similar measures.

In this analysis, we built a simulation model using National statistical data to record patterns of movements to work, and implement levels of mobility recorded in real time via mobile phone apps. This model was fitted to the pattern of deaths due to COVID-19 using approximate Bayesian inference. Our model is able to recapitulate mortality considering the number of deaths and datazones (DZs, which are areas containing approximately 500-1000 residents) with deaths, as measured across 32 individual council areas (CAs) in Scotland. Our model recreates a trajectory consistent with the observed data until 1^st^ of July. According to the model, most transmission was occurring “locally” (i.e. in the model, 80% of transmission events occurred within spatially defined “communities” of approximately 100 individuals). We show that the net effect of the various restrictions put into place in March can be captured by a reduction in transmission down to 12% of its pre-lockdown rate effective 28^th^ March. By comparing different approaches to reducing transmission, we show that, while the timing of COVID-19 restrictions influences the role of the transmission rate on the number of COVID-related deaths, early reduction in long distance movements does not reduce death rates significantly. As this movement of individuals from more infected areas to less infected areas has a minimal impact on transmission, this suggests that the fraction of population already immune in infected communities was not a significant factor in these early stages of the national epidemic even when local clustering of infection is taken into account.

The best fit model also shows a considerable influence of the health index of deprivation (part of the “index of multiple deprivations”) on mortality. The most likely value has the CA with the highest level of health-related deprivation to have on average, a 2.45 times greater mortality rate due to COVID-19 compared to the CA with the lowest, showing the impact of health-related deprivation even in the early stages of the COVID-19 national epidemic.

## Introduction

The COVID-19 pandemic was first introduced into Scotland no later than February 2020.^8^ Following a series of lesser restrictions and recommendations, on 23^rd^ March widespread restrictions were put in place in Scotland in accordance with the rest of the UK, with many businesses closed and most individuals except “key workers” restricted to movements only a very close distance from their homes. It is now known that this combination of measures has been highly effective in reducing the transmission of COVID-19 in Scotland such that the reproduction number fell below one by approximately the first week of April.^9^ However, as of yet there has been no study estimating the impact of different aspects of these control policies. Further, while there have been several studies showing substantial variation in COVID-19 mortality risk (Patel, et al., 2020), any assessment of the direct impact on mortality should consider the combination of transmission dynamics (influencing rate of spread and therefore the number of individuals exposed to infection) with deprivation related mortality. In this analysis, we use an explicitly spatial individual-based simulation model that accounts for recorded commuter movements, and geographically explicit population age structures, to estimate transmission characteristics by fitting a model to the observed number of COVID-19 related deaths in an Approximate Bayesian inference framework. Our aim is to estimate the impact of long distance travel restrictions and transmission reduction on the spread of COVID-19, and to assess the impact of spatially explicit measures of population health in order to estimate its impact on COVID-19 related mortality.

## 1. Model overview/methods

### 1.i Data sources

We use a combination of publicly available Scottish census data plus data on Scottish COVID-19 statistics provided by Public Health Scotland.

All statistics on COVID-19 deaths are drawn from records held by Public Health Scotland managed by the Electronic Data Research and Innovation Service or ‘eDRIS’ made available at the data zone (DZ) level where DZs are population census units of approximately 500 to 1000 residents (here, recorded in the 2011 census).^10^ We use publicly available census data on age demographics and movement to work patterns, available at the level of Census Output Areas (OAs), each of which contains approximately 20 households or 50 people.^11 12,13^. We also use the census data on the ‘Scottish index of multiple deprivation’ or SIMD. If an area is identified as ‘deprived’, this can relate to people having a low income but it can also mean fewer resources or opportunities. The SIMD is a relative measure of deprivation that considers multiple measures and combines them into a single value.^14^ Deprivation data are not publicly available at the OA level, but instead are recorded in DZs.^15^

To estimate reductions in mobility as a result of the restrictions of March 23^rd^, we use publicly available estimates by Google.^16^

### 1.ii. Compartmental model

The simulation framework (hereafter referred to as the Scotland Coronavirus Transmission Model, or SCoVMod) considers key aspects of COVID-19 epidemiology, including a latent phase, mildly infectious and highly infectious individuals, hospitalisation, recovery and death, similar to models used for other investigations (Arenas, et al., 2020; Di Domenico, Pullano, Sabbatini, Boëlle, & Colizza, 2020). These epidemiological processes are captured in a compartmental model (Figure 1). The model is stratified by three age groups: 0-15, 16-64 and 65+ years. We capture within OA transmission by assuming homogeneous mixing and between-OA transmission using population movement parameterised by empirical age-specific patterns of home and work contact. Individuals in infectious stages have the potential to infect those susceptible when co-located in the same OA at the same time (considering date and day/night patterns).

**Figure 1.**
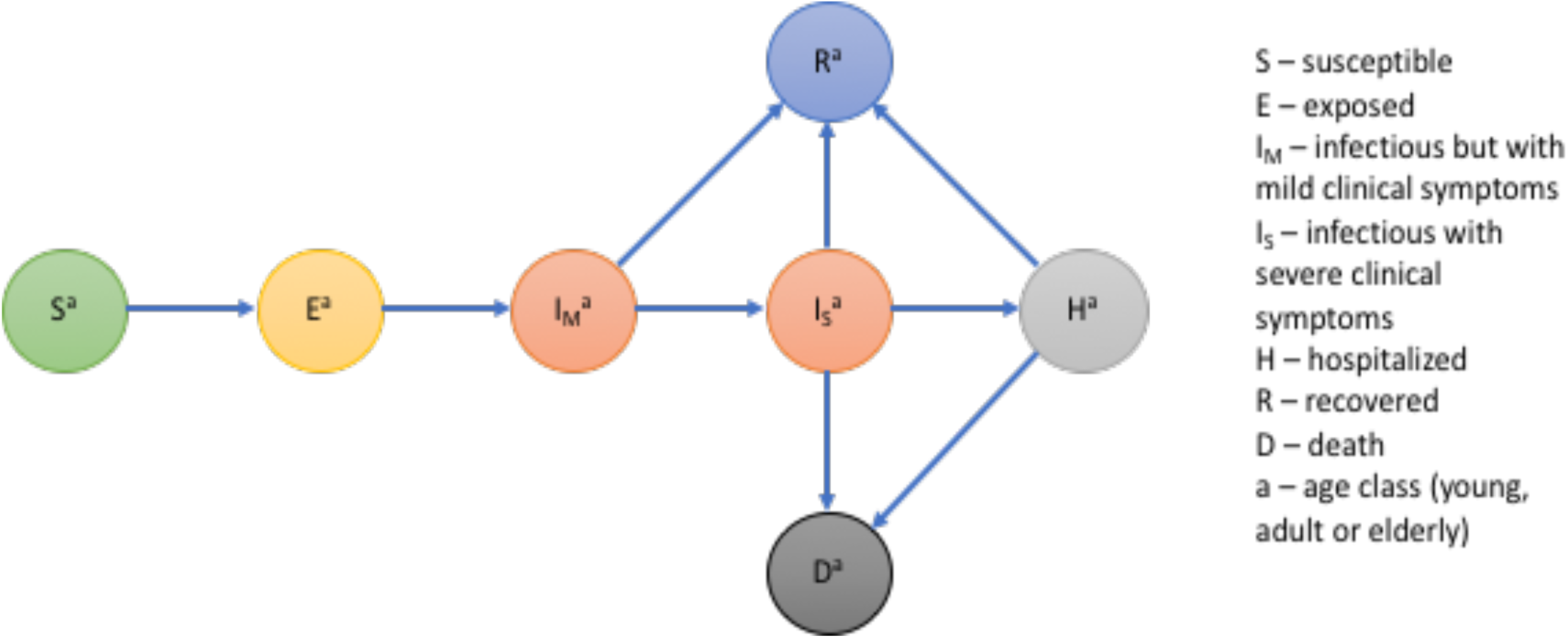
Schematic of infection stages in SCoVMod. Individuals pass through stages post infection as described by arrows. Not all stages are obligatory for all infected individuals (e.g. some individuals recover without going to hospital).

We assume that while the death rates for all age classes are the same, recovery rates differ, resulting in age-dependent differences across ages, in the proportion of individuals who die (see Table 1 for supporting references). Reports show that deprivation is an important indicator of COVID-19 mortality (Williamson, et al., 2020). Therefore, we consider the role of deprivation by adjusting for health index in the model (see Appendix I). Here, we use these factors to drive regionally specific differences, allowing for health index adjusted mortality rates by CA. It is assumed that the spatial patterns of infection are driven by commuter movements; i.e. movements between OAs lasting less than a day. We do not consider overnight shifts in location or introductions from outside Scotland beyond the impact on the initial seeding.

**Table 1.** Epidemiological parameters in SCoVMod, with priors and fixed values as appropriate. Where age is not indicated, parameters are assumed to be age independent.

| Parameters | Transiti<br>on | Symbol | Age | Estimate for<br>configured<br>parameters | Prior | Refs |
| --- | --- | --- | --- | --- | --- | --- |
| Latency period<br>(days) | $E \rightarrow I_M$ | $1/\gamma$ | All | *fitted* | $U(1.67, 2.8)$ | (Arenas, et al., 2020; He, et al., 2020; Li, et al., 2020; Bi, et al., 2020; Zhang, et al., 2020) |
| Time from mild infectiousness to recovery (days) | $I_M \rightarrow R$ | $1/\rho_M$ | All | *fitted* | $U(0.67, 2.8)$ | (Di Domenico, Pullano, Sabbatini, Boëlle, & Colizza, 2020; Zhang, et al., 2020) |
| Symptoms onset time after infectiousness (i.e. incubation period – latency period) (days) | $I_M \rightarrow I_S$ | $1/\gamma_M$ | All | *fitted* | $U(2, 28)$ | (Arenas, et al., 2020; He, et al., 2020; Li, et al., 2020; Bi, et al., 2020; Zhang, et al., 2020; Linton, et al., 2020; Lauer, et al., 2020; Sanche, et al., High Contagiousness and Rapid Spread of Severe Acute Respiratory Syndrome Coronavirus 2, 2020; Sanche, et al., The novel coronavirus, 2019-nCoV, is highly contagious and more infectious than initially estimated, 2020) |
| Transmission rate for severe infectors $I_S$ (baseline, daytime) | $S \rightarrow E$ | $\beta_d$ | All | *fitted* | $U(0, 2.8)$ | - |
| Transmission rate for severe infectors $I_S$ (baseline, nighttime) | $S \rightarrow E$ | $\beta_N$ | All | *fitted* | $U(0, 2.6)$ | - |
| Transmission rate multiplier for mild infectors $I_M$ | $S \rightarrow E$ | $\gamma$ | All | *fitted* | $U(0, 2.6)$ | - |
| Time from severe symptom onset to hospitalization (days) | $I_S \rightarrow H$ | $1/\eta$ | All | 4.00 | - | (Sanche, et al., The novel coronavirus, 2019-nCoV, is highly contagious and more infectious than initially estimated, 2020; Sanche, et al., High Contagiousness and Rapid Spread of Severe Acute Respiratory Syndrome Coronavirus 2, 2020; Qin, et al., 2020; Wang, et al., 2020; Liu, et al., 2020; nan Han, et al., 2020; Pung, et al., 2020; Verity, et al., 2020; Filipe, et al., 2020) |
| Time from severe symptom onset to recovery for non-hospitalised (days) | $I_S \rightarrow R$ | $1/\rho_S$ | Young | 19.00 | - | (Sanche, et al., The novel coronavirus, 2019-nCoV, is highly contagious and more infectious than initially estimated, 2020; Arentz, et al., 2020) |
|  |  |  | Adults | 20.70 |  |  |
|  |  |  | Elderly | 21.60 |  |  |
| Time from hospitalization to death (days) | $H \rightarrow D$ | $1/\mu_H$ | Young | - | - | eDRIS data |
|  |  |  | Adults | 6.97 |  | eDRIS data |
|  |  |  | Elderly | 6.62 |  | eDRIS data |
| Proportion of hospitalized who recover | $H \rightarrow R$ | $\rho_H/(\rho_H + \mu_H)$ | Young | 1.00 | - | eDRIS data |
|  |  |  | Adults | 0.96 |  | eDRIS data |
|  |  |  | Elderly | 0.84 |  | eDRIS data |
| Time from symptoms onset to death (days) | $I_S \rightarrow D$ | $1/\mu_S$ | Adults | 16.00 | - | (Sanche, et al., High Contagiousness and Rapid Spread of Severe Acute Respiratory Syndrome Coronavirus 2, 2020; Qin, et al., 2020; Liu, et al., 2020; Arentz, et al., 2020; Filipe, et al., 2020; Verity, et al., 2020) |
| mortality rate multiplier (relative to average health index) | | $\mu_{mod}$ | All | *fitted* | $U(0, 0.08)$ | |
| Number of seed infections | | $N_s$ | N/A | *fitted* | $U(0, 2000)$ | |

Because our initial analysis concentrates on the early weeks of the epidemic dominated by pre-lockdown infection spread, we make a number of simplifying assumptions regarding transmission pathways: first we do not model infections in care homes as they are assumed to result in few additional infections outside of these locations. Similarly we assume hospital-acquired infections do not result in substantial community outbreaks (Treibel, et al., 2020). We assume that only adults contribute to commuter movement; in the daytime, the remaining proportion of adults and all young and elderly individuals are assumed to move primarily within their local OAs, which also account for non-work activities. Commuting is also restricted to healthy and exposed or mildly symptomatic individuals; severe infections and hospitalised individuals do not commute.

The equations for the complete model are provided in Appendix II.

### 1.iii Modelling Lockdown

We model the impact of lockdown using two effects. First, we model the reduction in longer distance activity by reducing the volume of commuter activity or the “mobility-reduction” scenario. We thin the number of movements to be consistent with the observed decline mobility, as measured through Google mobility data for Scotland ^17^ and social contact surveys (Jarvis, et al., 2020). We compared this assumption to an independent dataset to corroborate the reduction in rural areas, where data are few (see appendix V). Second, we model physical distancing measures as a reduction in the contacts (including both within the OA of residence, and the daytime OA that includes work locations). This directly influences transmission rates, therefore we call this the “transmission-reduction” scenario. For the post-lockdown period, we fit a reduction in the transmission rate independently of the other parameters assuming that the posterior distributions from the pre-lockdown fit (for infectious periods, mortality rates etc.) remain relevant to the post-lockdown period. We therefore vary only the effective date of reduction in transmission, and its amount, considering a range of values that are broadly consistent with the observed reduction in the effective reproduction number “R^t^”^18^.

### 1.iv Influence of Health Index on Mortality

Based on NRS data on Covid-19 related deaths that were outside care homes, we examined the impact of different deprivation factors relevant to this period. We found that whilst deprivation overall (as measured by the SIMD) is significantly associated with increased Covid-19 mortality. This was further disaggregated as follows (see also appendix I):

1. Population level risk of Covid-19 mortality is associated with the SIMD indicator that describes (good) accessibility and orthogonally with the SIMD indicator that describes (poor) health, indicating that areas with poorest health and good access experienced higher Covid-19 mortality.
2. Risk of excess Covid-19 mortality (Covid-19 deaths as a fraction of all deaths) is most closely associated with the access indicator component of SIMD. This indicates that the areas that have good local connectivity and transport will have higher rates of Covid-19 transmission.

We hypothesize that “access” is a proxy for transmission model dynamics in two ways; first, transmission rates in the model may be influenced by access (with greater access likely implying a higher probability of introduction of infection and therefore earlier introduction. Thus the observed differences in mortality overall is dependent on the time since introduction (and therefore the initial seeding plus transmission dynamics), and health. We therefore fit a modifier to the COVID-19 mortality rate in our model using the health index (only) considering the average health index in each CA:

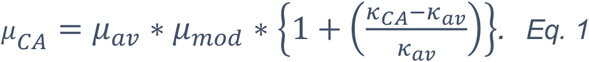

Here *μ*_*CA*_ is the COVID-19 related mortality rate for a given CA, *μ*_*av*_ Is the average across CAs, *k*_*CA*_ a is the CA mean health index value (from the SIMD), and “*μ*_*mod*_” is a fitted parameter given a prior range of [0, 0.08] in order to precludes negative values for low values of *κ*_*CA*_.

### 1.v Model Inference and Computation

The population structure, movement patterns (see appendix IV for details) and the infection model are used to generate simulated epidemics which iterate over half-day increments to consider two types of contact: non-commuting/home locations, and locations where commuting individuals interact. Homogeneous mixing at the OA level is assumed. Simulated epidemics are compared in space and time to the recorded pattern of COVID-19 spread in Scotland.

All non-observable or unknown parameters were estimated using the number of recorded deaths due to COVID-19 related causes, considering all weeks beginning 9^th^ March and ending on the 12^th^ April 2020. Recorded deaths are from the weekly NRS records with COVID-19 related causes, identified by the DZ of residence.^19^ Especially in the early stages of the epidemic these are the most complete and unbiased indicators of infection available. However, they also differ slightly from other official sources which record the date of death, rather than the date of registration of death. We assume that for each reported case, death occurs in the week prior to the registered week as registration is allowed up to 8 days post mortem. Estimation was performed using a sequential Monte Carlo implementation of Approximate Bayesian Computation (ABC-SMC) (Hartig, Calabrese, Reineking, Wiegand, & Huth, 2011; Toni, Welch, Strelkowa, Ipsen, & Stumpf, 2009). Specifically, we calibrated the model output to the cumulative weekly number of deaths due to COVID-19 aggregated at the level of local government areas (“Council Areas” or CAs).^20^ Preliminary attempts to fit the data using weekly incidence proved to have poor results. We hypothesized that this was a result of a combination of very few deaths to constrain the fits in the early stages, and also the complications from the multiple changes in the control efforts especially through March. Thus we chose to emphasize the latter stages of the period by choosing as our observation the cumulative number of all COVID-19 related deaths per Council Area. The metric used to compare simulated and observed summary statistics was defined as a sum of squared errors (SSE) of this number, recorded weekly:

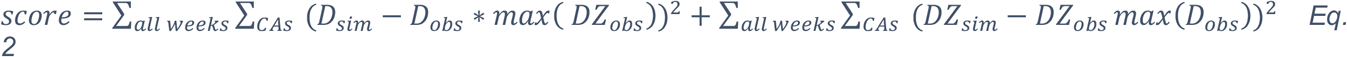

Where:

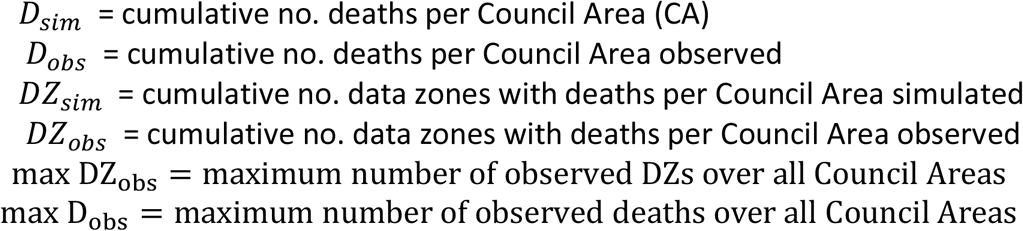

Uniform prior distributions were applied to parameters to constrain their values to plausible ranges based on the available literature relevant to the early, pre-lockdown period (Table 1). Infection dynamics are simulated via a τ-leap algorithm using half-day timesteps (Gillespie D. T., 2001). All parameters are listed in Table 1.

### 1.vi Model Seeding

The total number of infected individuals at the start of the simulation (the “seeds”) are fitted as part of the inference. The seeds are randomly assigned a disease state from *E, I*_*M*_, and *I*_*S*_. To establish an approximately representative spatial distribution, seed locations are also assigned randomly, according to the cumulative proportion of deaths registered per ‘intermediate’ zone, as recorded up to the week of March 23^rd^, 2020. Intermediate zones are aggregates of approximately five Data zones; this unit is chosen for seeding to account for clustering of infections in areas near to identified deaths.

## 2. Results

### 2.i Movement and network patterns

The patterns of movement generated at the OA level, shows some substantial differences in both the average distance travelled, and the connectedness between OAs across the country. Predictiably, individuals in rural areas seem to move the farthest to work on average, and individuals in the densely populated ‘Central Belt’ are the most connected (Figure 2).

**Figure 2.**
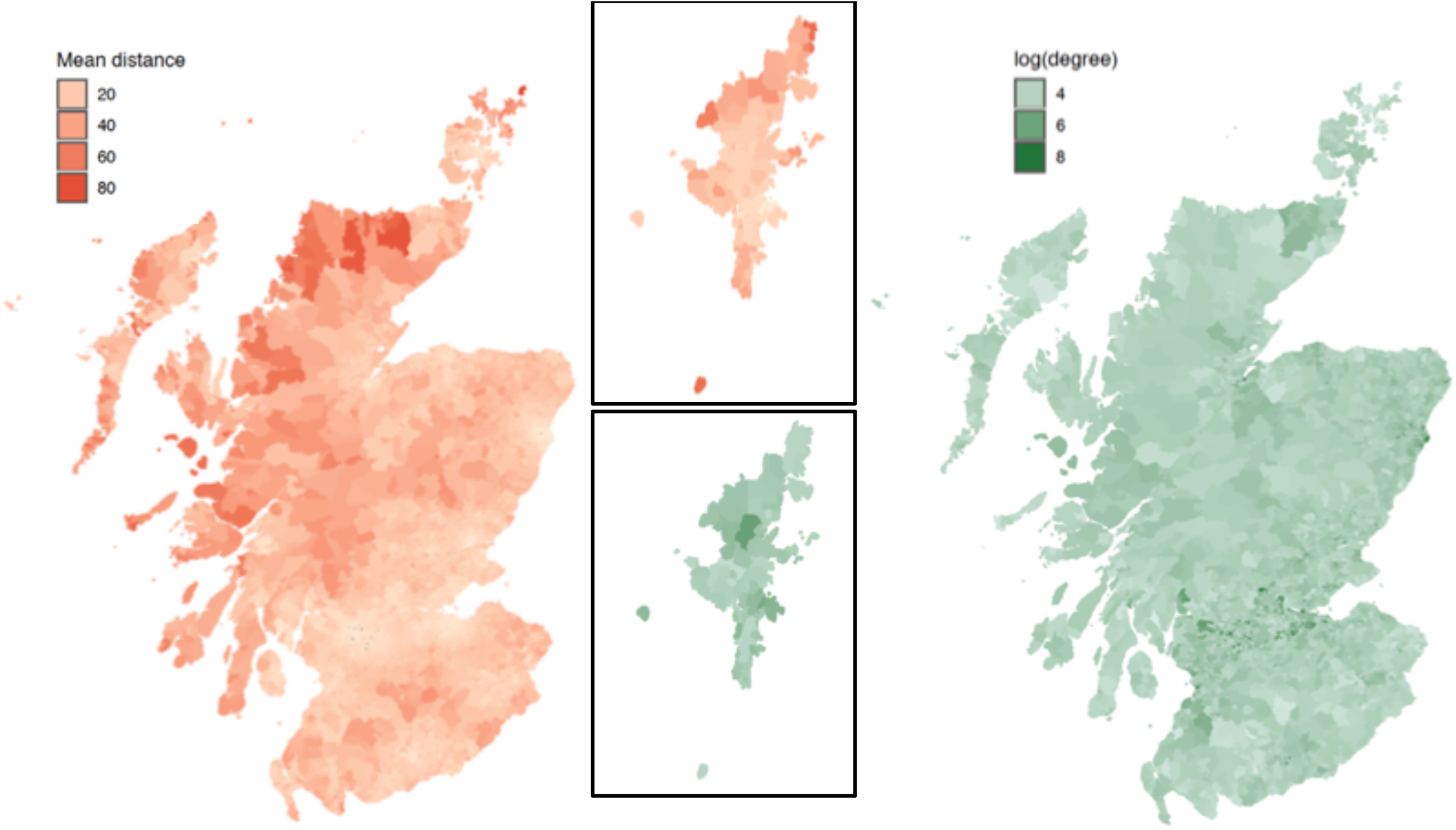
Commuter movement patterns, with commuters aggregated by output area (OA; Census areas with typically 50- 500 individuals; maximum of 2081). Data according to the 2011 census. (L) mean distance travelled from OA in km. and (R) mean number of OA’s to which each OA is connected to. The greatest distances are travelled on average by individuals in remote locations. The greatest network degree is found in highly urbanised areas

### 2.ii Parameter Posteriors and Model Fit

Posterior parameter distributions are strongly uni-modal across all parameters with no evidence of strong correlations across any pair of parameters [show in appendix VI]. Of particular interest is the increase in death rate due to deprivation. The most likely estimate for the modifier to mortality is *μ*_*mod*_ = 0.03, resulting in a difference in the death rate of 2.45 times comparing the highest to the lowest health index CAs. Post-lockdown, the best fit value (lowest metric value of Equation 1) is found to occur when transmission rates are reduced to 0.12 times the value, as of 28^th^ March 2020. In figure 3, the fitted model is shown to reproduce the epidemic curve over the period from beginning of March to July 1^st^, with the number of observed deaths per CA lying within the range of 95% of the fitted, simulated epidemics (parameters drawn as sets from the posterior parameter distributions). The number of DZs with deaths more often exceeds these limits (figure 4) however we note that the low numbers of such zones per week, makes the impact of stochastic variability higher. Where the number of DZs with deaths is higher (e.g. the Cities of Edinburgh and Glasgow), fidelity is generally greater.

**Figure 3.**
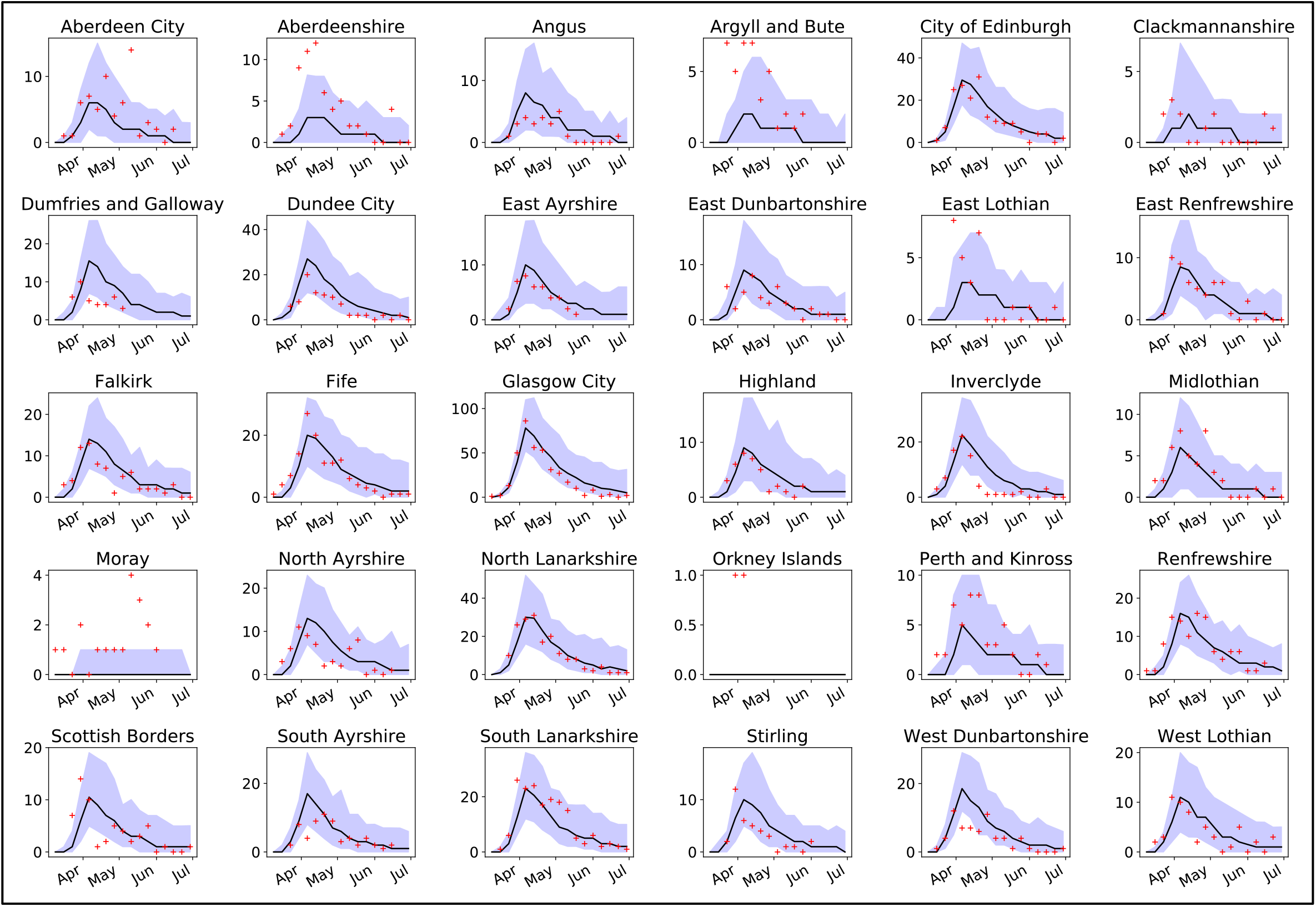
Number of deaths per week for all Local Council Areas in Scotland (bar Shetland). Incidence number (red crosses) of COVID-19 related deaths (upper plot) compared to the median of 100 simulations (black line), and 95% C.I.’s (purple areas).

**Figure 4.**
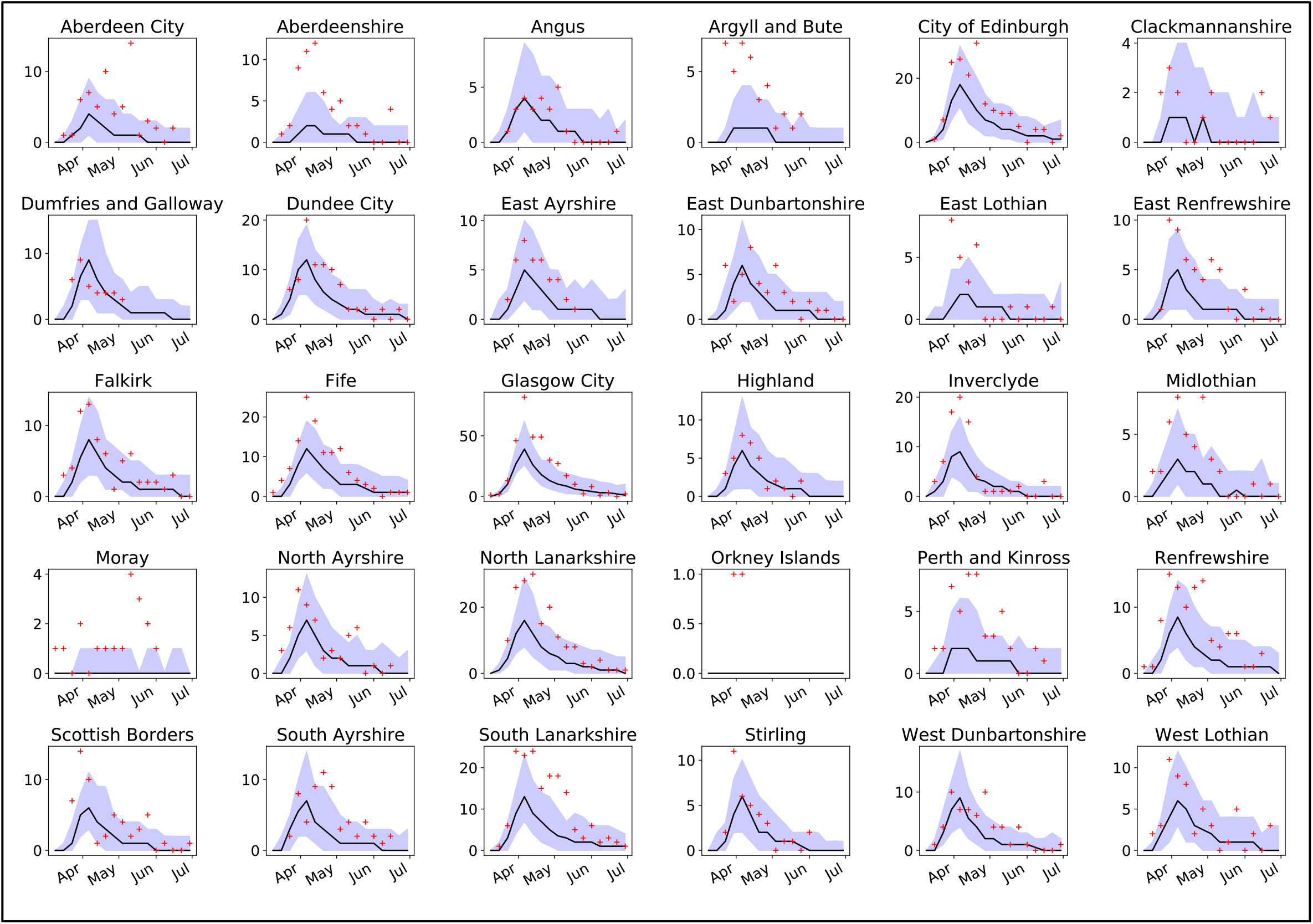
Number of data zones (DZ’s) with deaths per week for all Local Council Areas in Scotland (bar Shetland). Incidence number (red crosses) of COVID-19 related DZ’s with deaths (upper plot) compared to the median of 100 simulations (black line), and 95% C.I.’s (purple areas).

The best fit model shows a considerable influence of health index on mortality; the most likely impact is that the CA with the worst index will have, on average, an increase in mortality of approximately 2.45x compared to the CA with the worst health index (cf. Equation [2]).

### 2.iii impact of distance reduction and transmission reduction on COVID-19 spread

Lockdown restrictions are likely to impact both the local spread of infection (via social distancing measures and therefore change the transmission rates *β*_*D*_ and *β*_*N*_) and the spatial spread of infection (e.g. limitations on travel by car). Most infection pressure occurs within OA, but with a few OAs where commuting traffic produces a strong spatial signature (figure 5a); these are typically urban areas, most likely with considerable inward commuting traffic (Figure 5b). In most areas, however, disease spread is predominantly local, with the most likely outcome being approximately 90% of infection occurring within OA. We illustrate the impact of lockdown on spatial spread, by considering the likely extent of disease should lockdown restrictions have been imposed two weeks prior to the actual date (23^rd^ March 2020) which is approximately the time of the announcement of the first death due to COVID-19 in Scotland, and compare scenarios where only the transmission rates (determined by changes in local activities such as via the 2m rule limiting distance of contact) are reduced, or only the commuting fraction is reduced (restrictions to movements close to home) or both are reduced (i.e. lockdown as it occurred but two weeks earlier). With earlier lockdown, we predict on average 581 deaths (95% of simulations within 377 to 1010 deaths) by 26^th^ April 2020, compared to 2722 (range 1294 to 4050) in the baseline scenario (observed number is 2,795, assuming that all deaths occur in the week prior to the week the death is registered in) (figure 7).

**Figure 5a.**
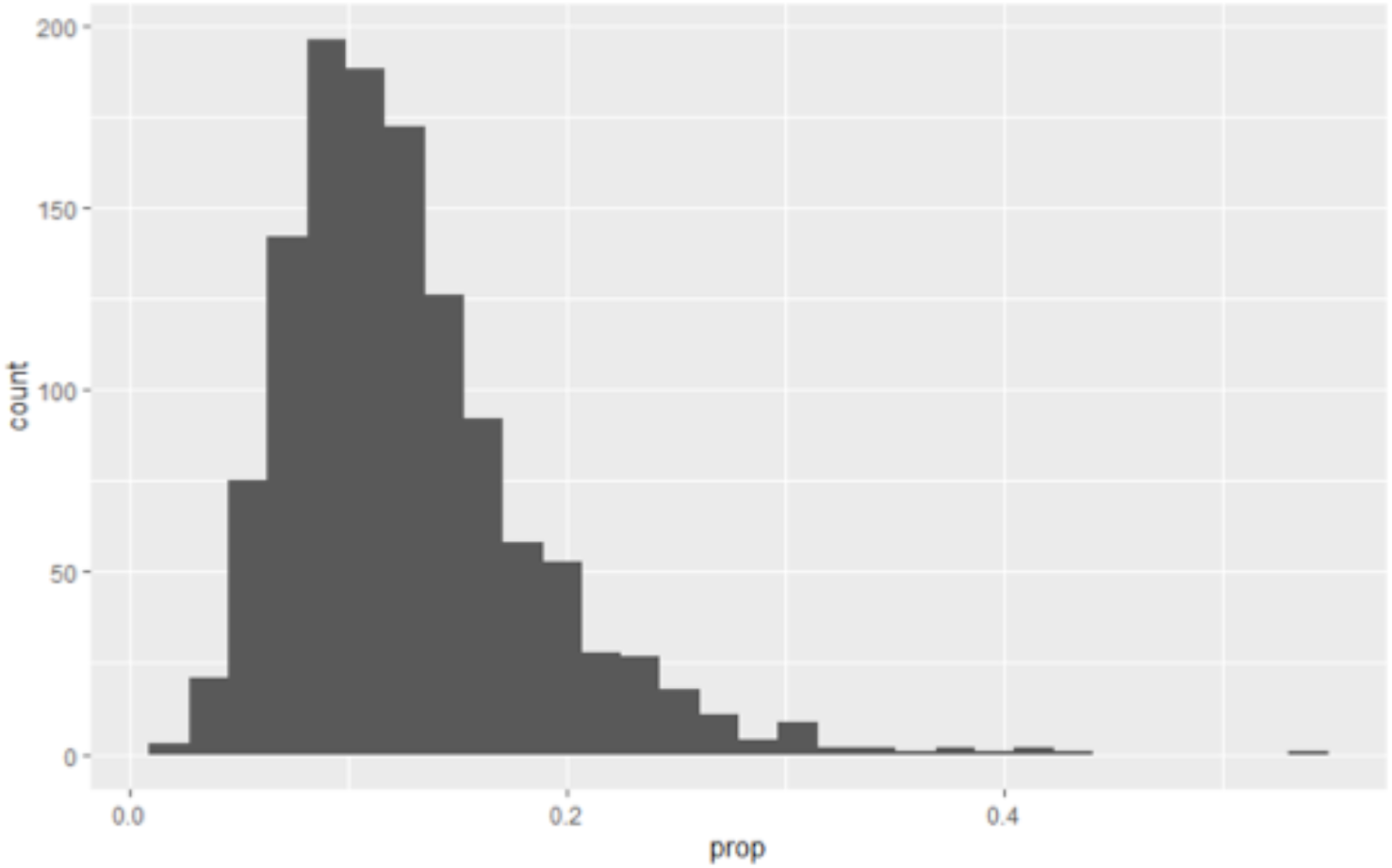
Proportion of transmission occurring outside of OA’s of residence, across all OA’s in Scotland. For most OA’s the majority of transmission is estimated to occur within the home location.

**Figure 5b.**
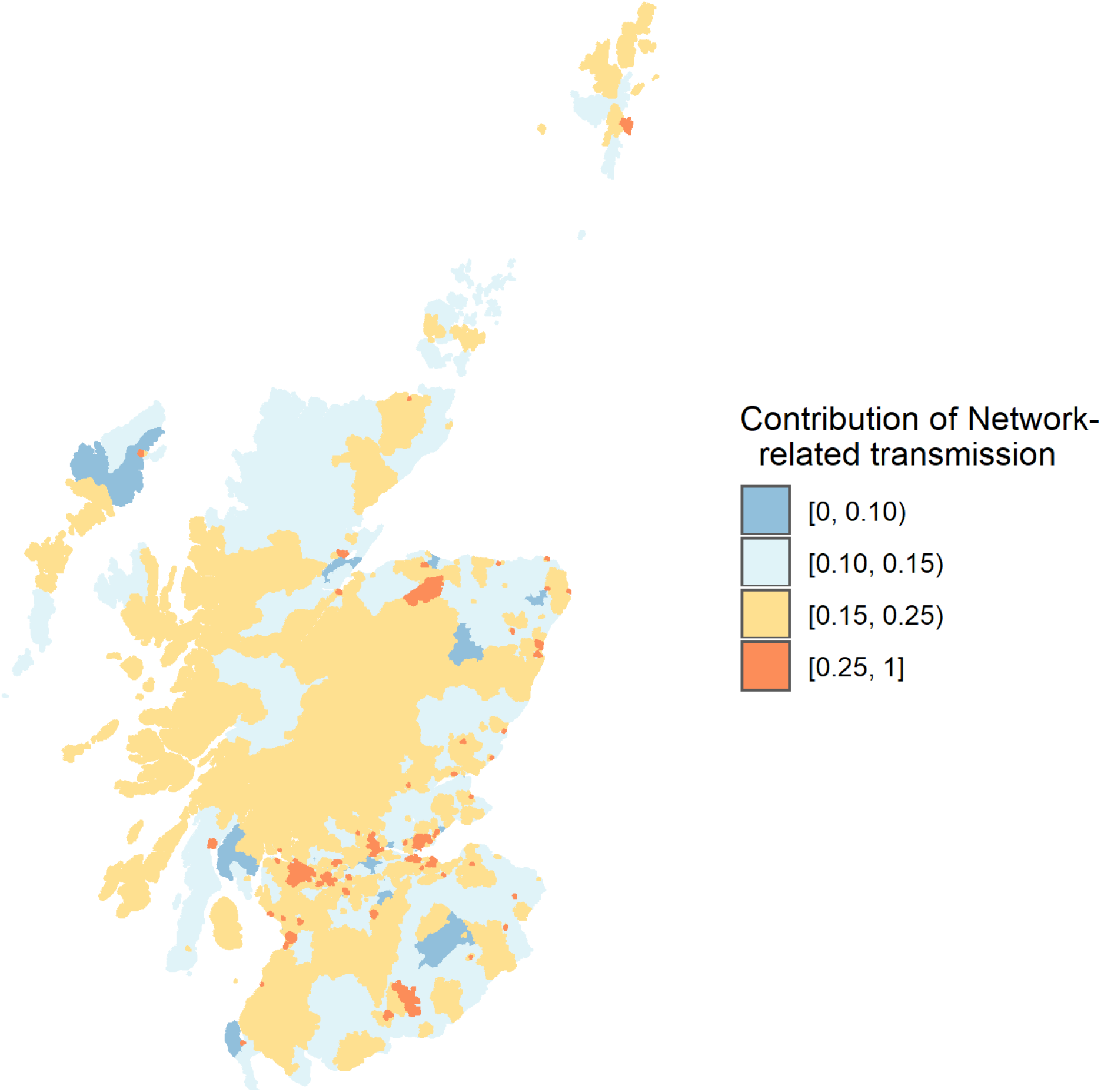
Estimated relative proportion of the transmission potential that is within OA and outside of OA. Areas in yellow (high) and orange (extremely high) represent highly urban areas, with considerable inward commuting traffic.

**Figure 6.**
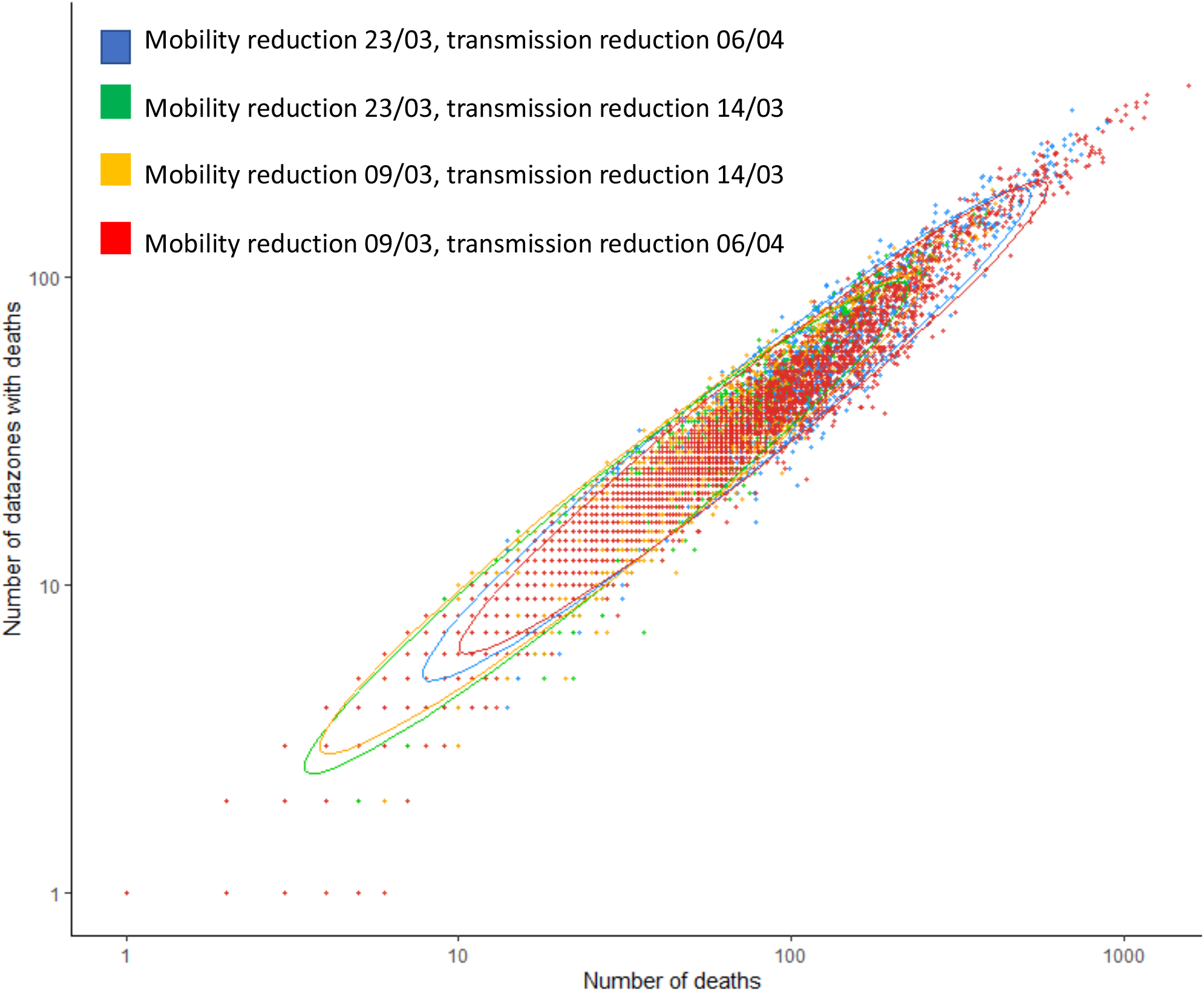
Comparison of number of COVID-19 related deaths and number of datazones affected as of April 26^th^ 2020, contrasting baseline (imposition of restrictions as they occurred), early (March 9^th^) imposition of social distancing measures but without restriction of long distance travel (early beta), early imposition of long distance travel restriction only (early lockdown) and early imposition of both.

**Figure 7.**
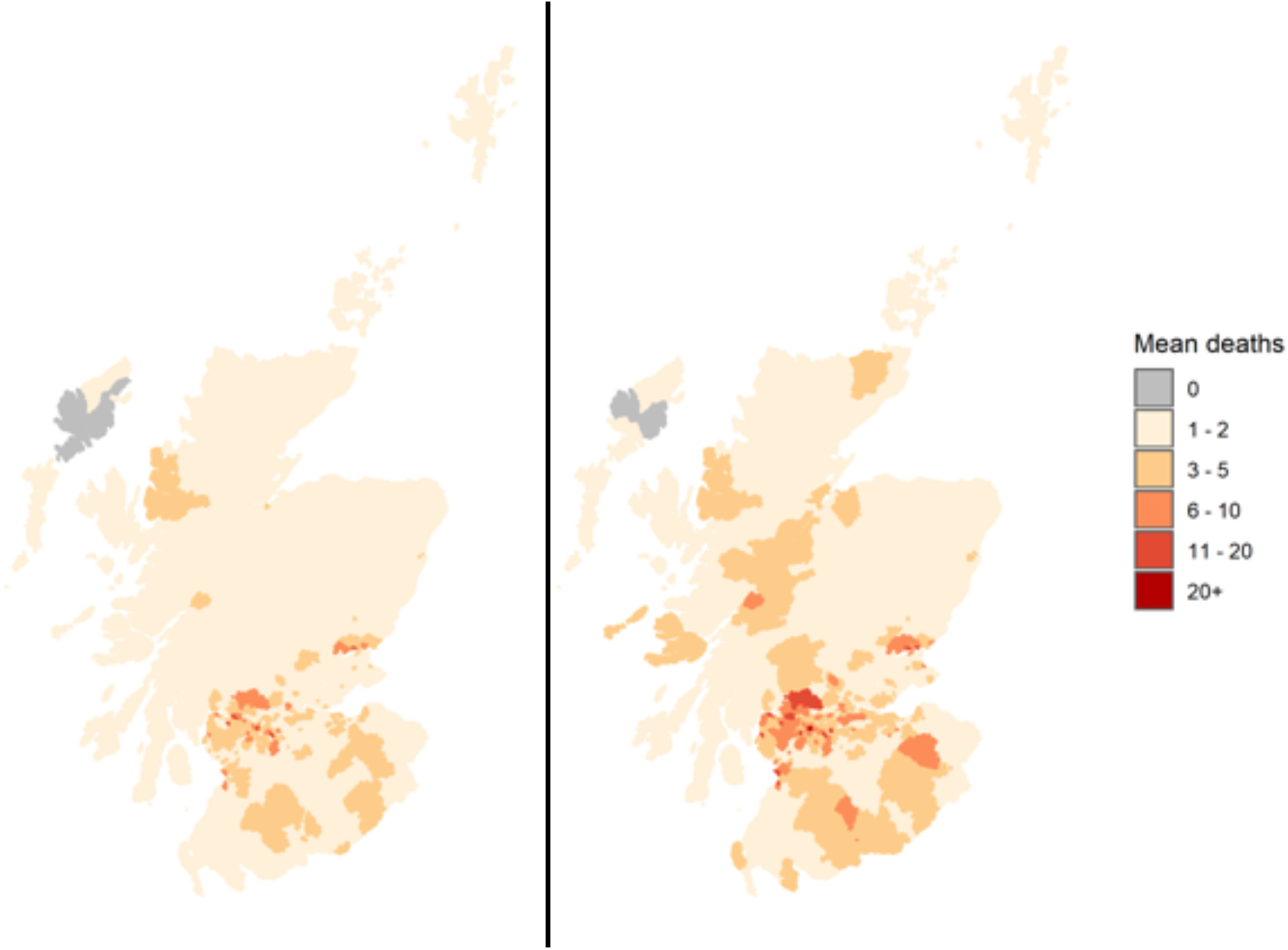
Mean number of deaths per OA, averaged over 50 simulations for early lockdown on 16^th^ March 2020 (L) and lockdown as it occurred on 23^rd^ March 2020 (R) considering the total number to 30^th^ April, 2020.

The reduction in the total number of deaths also results in a reduction in geographical spread with many fewer DZs affected by COVID-19 mortality in the early lockdown scenario (figure 7).

## 3. Discussion

In this report we present the first description of spatio-temporal dynamics of COVID-19 in Scotland. Exploiting our estimates of the relative contribution of long distance transmission (driven by to-work movements) and local transmission (including work and non-work interactions) we compare different timings of elements of the control policy, separating out the restrictions on non-essential work activity from the social distancing that would have influenced the transmission rate. Our model outputs show good fidelity to both the number of deaths and a measure of the spatial spread of observed deaths, providing confidence that we have captured some essential features of the epidemic in Scotland. We show that at this early stage, social distancing was dominant in reducing the death rate, with reduction in movement distances having little impact on mortality. We suggest that the impact of reducing long distance travel alone is small because individuals who are restricted from long distance travel instead continue to transmit in their ‘home’ locations. This imposes an additional infection pressure in their local OA and in the absence of substantial levels of immunity to limit transmission, results in similar numbers of new infections. Were this not to be the case, increasing the number of movements out of OA would serve to increase the probability of moving infected individuals to immune-naïve locations. Unless there is an inherently lower transmission risk in these immune-naïve locations the expectation is that this translocation would likely increase the overall number of infections, and therefore deaths. The combination of long distance restrictions and transmission reduction is considerably more potent, with substantial reductions in both the areas being affected (allowing for potentially more targeted control) and total deaths (both directly saving lives, and reducing impact on the health care system).

The fitted parameters suggest that mildly infected individuals play an important role in transmission. This does not necessarily imply higher levels of infectiousness compared to severely infected, as it is the mildly infected individuals who are assumed to be responsible for long distance spread (i.e. that severely infected individuals would likely stay at home). Our model also estimates the number of individuals with infection in early March, with the most likely case being on the order of 1500 infected individuals in Scotland at this time. While this approach is crude (it does not take account for the importation of individuals over time), it is substantially greater than the estimate of 113 introductions of COVID-19 into Scotland based on viral sequence data, but plausible if one takes into account the additional infections these introductions would have caused by the time of our simulation start in the early pandemic period when there were no local restrictions in place.

An important cautionary note is that this assessment does not consider the longer term impact of reducing geographical spread and so should not devalue the impact of long distance movement restrictions. More localised ‘clustered’ infections ultimately result in a higher proportion of recovered individuals in ‘hotspot’ areas (promoting localised herd immunity effects). Further, as these hotspots were typically in more urban areas (likely caused by the higher number of movements into Scotland from previously affected areas), this also has the important consideration of reducing infection rates in remote areas, where access to hospitals and ICU is typically poor. Our presumed patterns of movements are based on movements to work – a considerable portion of activity is of course non-work related and so our assumption is therefore that these patterns do not deviate considerably in terms of the spatial distance associated with movement to work patterns.

By explicitly modelling transmission dynamics, this allows us with relatively few data, to infer in our model that health-related deprivation results in an 2.45x (mostly likely value using equation 2) difference in the death rate due to COVID-19 between the council area with the highest average health index and that with the lowest. Deprivation (including health) in Scotland varies substantially at a much finer scale, with data zones with the highest deprivation often neighbouring zones with the lowest. This suggests that, were the data available, a deeper interrogation could provide a much more refined assessment of potential health burdens and risks associated with geographical spread.

The direct interpretation of the lockdown scenarios and in particular future projections must be viewed with some caution. The effectiveness of lockdown will vary in space and time, due to differences in human behaviour as has been seen elsewhere.^21^ On the other hand, reduced numbers of cases will reduce spread, and therefore logistical burden, with possibly more resources available and the burden on care homes, hospitals and ICUs reduced. These potentially counterbalancing factors would of course have to be considered in more detail. Despite these caveats, our simple approach is useful to strategically examine trade-offs between travel related restrictions, and social distancing when evaluating future releases from lockdown. “Small world” effects suggest that even a small number of long distance movements, joining up communities, are sufficient to cause widespread epidemics. Here however, most of the protection, even geographically is a result of transmission reduction; long distance restrictions only have a minimal impact most likely because in our model, transmission that does not occur at long distance does occur at the OA level, increasing the infection pressure from remaining commuters. Thus only extreme travel restrictions are likely to have an impact, at least until build-up of immunity levels is more substantial than observed here. The estimates of mobility under even extreme lockdown we use here suggest this kind of elimination is difficult to achieve, and thus any travel restrictions must be accompanied by measures to continue to reduce transmission if, in future, we are to prevent spread of COVID-19 to areas that have successfully eradicated local COVID-19 cases.

## Data Availability

All data used in this study are freely available (though in some cases subject to signing an appropriate data sharing agreement). Links to sources where these can be found are in the manuscript.

## Acknowledgements

Samantha J. Lycett (both Roslin Institute, University of Edinburgh), for helpful advice on the model development. Prof. C. Robertson (Strathclyde University, Health Protection Scotland) for advice on scenarios. Funding notes: this work has been funded by Roslin ISP2 (theme 3) - BBS/E/D/20002174, Wellcome Trust grant 209818/Z/17/Z, BBSRC grant BB/P010598/1, and Strategic Blue Cloud Fund to Fight COVID-19 grant 003

## Appendix I. Analysing deprivation indices in relation to Covid-19 data for Scotland

### Methods

The SIMD is based on a set of indicators at the data zone level (approximately 500 to 1000 persons per zone). The SIMD was estimated in 2016 and again in 2020, however, only the ranks are published for 2020 and we have been unable to access the methodology for estimating the raw SIMD scores for SIMD 2020. We therefore analyse here the 2016 data. The indicators that comprise the SIMD are broken down in Table 1.

**Table A.III.1.**
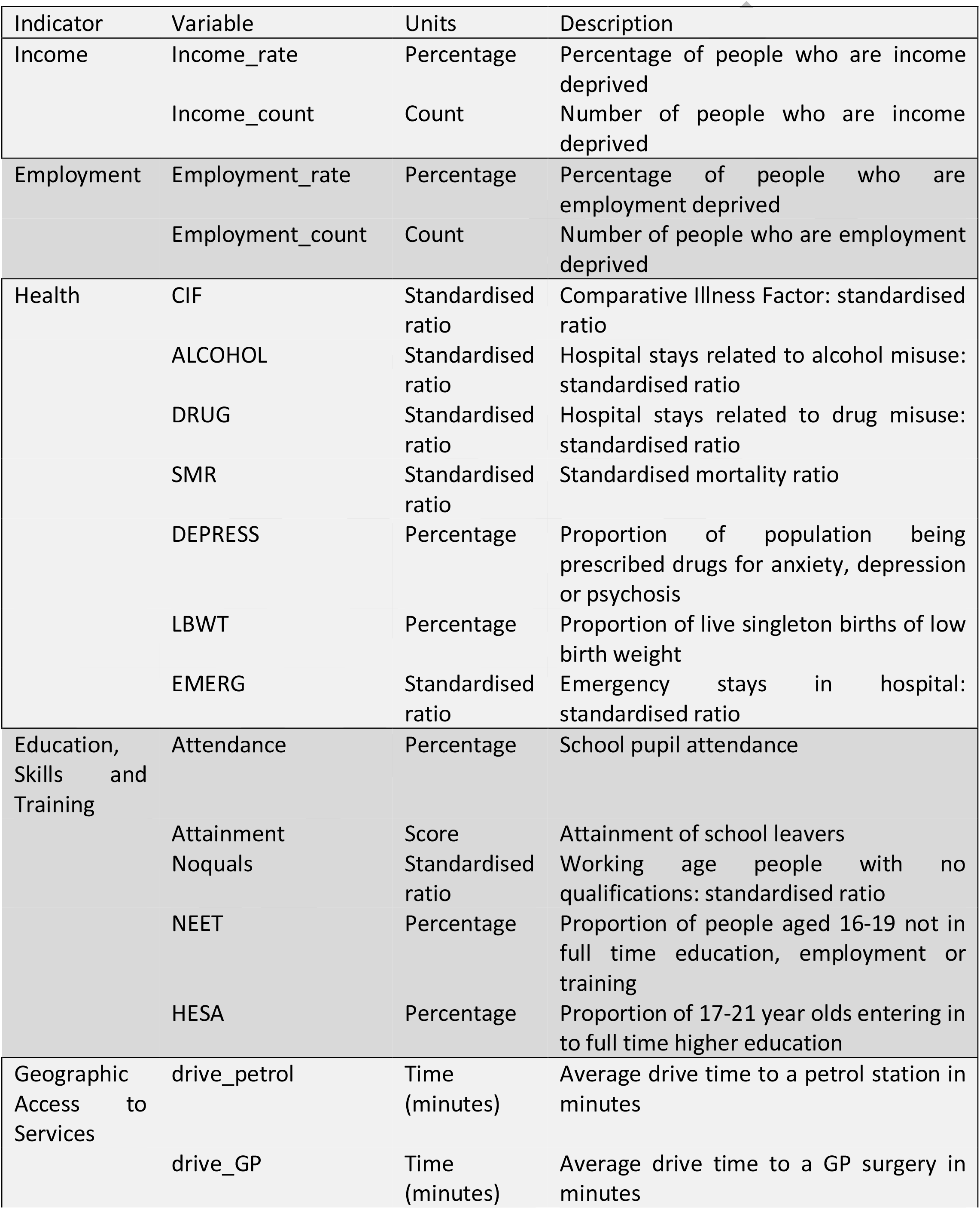
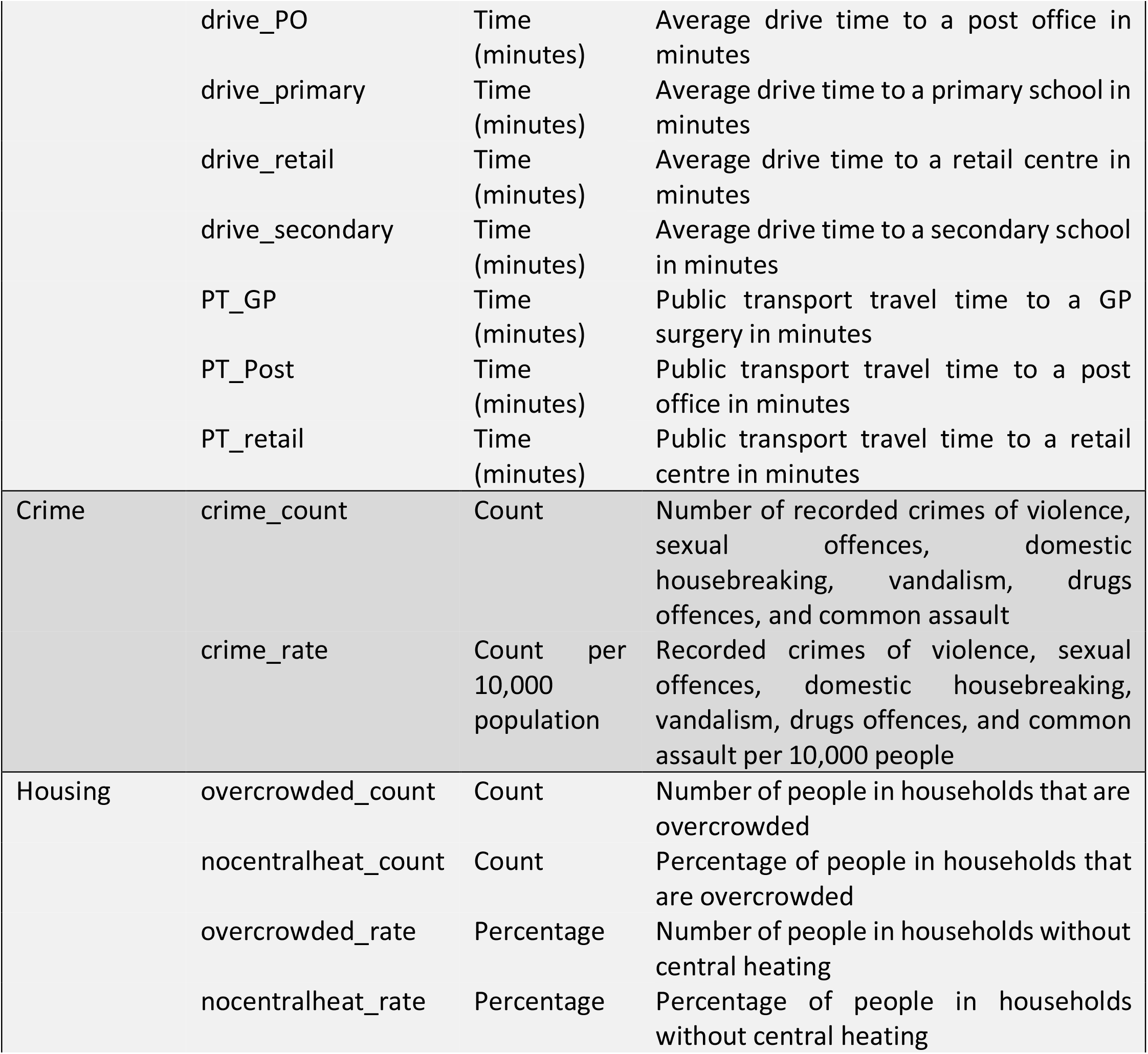
Indicators and variables that are used for estimating the SIMD 2016.

### Specific data were sourced from

1. Data on Covid-19 related deaths that were outside care homes were processed to the council area
2. Data on SIMD for 2016 and 2016 were downloaded from https://www.gov.scot/collections/scottish-index-of-multiple-deprivation-2020/ which provided ranks for data zones and each indicator at the data zone level
3. Raw data for SIMD 2016 were accessed through a GitHub repository (https://github.com/TheDataLabScotland/openSIMD). This repository included the weightings and methodology for calculating the individual indicator scores and the overall SIMD score rather than ranks alone.

### Statistical analysis

We constructed two models:

1. A population level mortality model, and
2. An excess deaths model.

The models were generalised linear models with binomial error structures at the local council level (N = 32). For the population level model the outcome variable was c(Covid deaths, population – Covid deaths). For the excess mortality model c(Covid deaths, all deaths – Covid deaths). To correct for overdispersion we fitted the model with an individual level random effect for each data point using the glmer function in the lme4 (1) package for R, in multivariable models variance inflation factors were checked using the car package (2) and overdispersion using the DHARMa package (3). We also checked the results against a model fitted using the quasi-binomial family. In both models we tested the following covariates in univariable analysis:

- Population (form SIMD 2020)
- Population density
- SIMD 2016 score
- SIMD 2016 score without the access component
- SIMD 2016 income indicator
- SIMD 2016 employment indicator
- SIMD 2016 health indicator
- SIMD 2016 education indicator
- SIMD 2016 housing indicator
- SIMD 2016 access indicator
- SIMD 2016 crime indicator

### Results

The results of univariable analyses of these variables for the population level is presented in Table A.III.2.

**Table A.III.2.** Results of the univariable model with Covid deaths / population as the outcome

| Predictor | Estimate | St. error | p-value | AIC | BIC |
| --- | --- | --- | --- | --- | --- |
| Population / 100000 | 0.0137 | 0.067 | 1.063 | 303.2 | 307.6 |
| SIMD score | 0.046 | 0.012 | <0.001 | 291.5 | 295.9 |
| SIMD education indicator | 0.036 | 0.013 | 0.005 | 297.0 | 301.4 |
| SIMD health indicator | 0.042 | 0.008 | <0.001 | 283.1 | 287.5 |
| SIMD housing indicator | 0.025 | 0.009 | 0.006 | 297.4 | 301.8 |
| SIMD access indicator | -0.040 | 0.009 | <0.001 | 283.8 | 288.2 |
| SIMD crime indicator | 0.049 | 0.013 | <0.001 | 290.7 | 295.1 |
| SIMD income indicator | 0.040 | 0.009 | <0.001 | 289.8 | 294.2 |
| SIMD employment indicator | 0.041 | 0.009 | <0.001 | 287.2 | 291.6 |
| SIMD score (exc access) | 0.047 | 0.011 | <0.001 | 288.2 | 292.6 |
| Sqrt(population density) | 0.017 | 0.006 | 0.003 | 295.9 | 300.3 |

As access has the lowest AIC and BIC and is orthogonal to the other variables we tested it against the remaining variables in turn to check for a multivariable model. The model with access and health had the lowest AIC (276.3), BIC (282.2) and low variance inflation factor (1.291) and was not overdispersed (Table AIII.3). The data are presented in Figure A.I.1.

**Table A.III.3.**
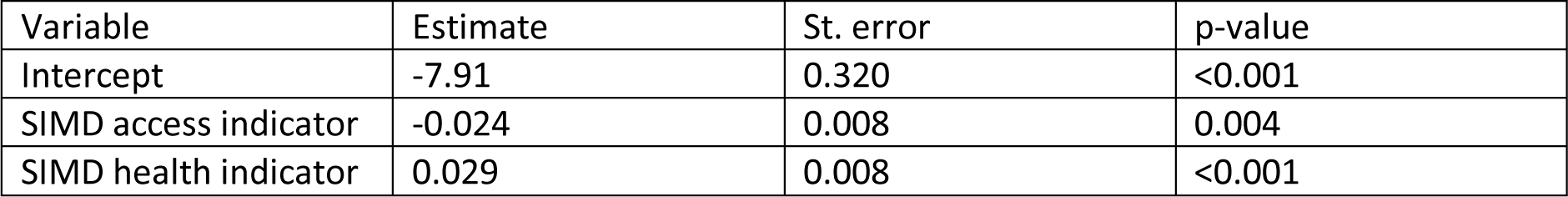
Results of the multivariable model with Covid deaths / population as the outcome

**Figure A.1.i.**
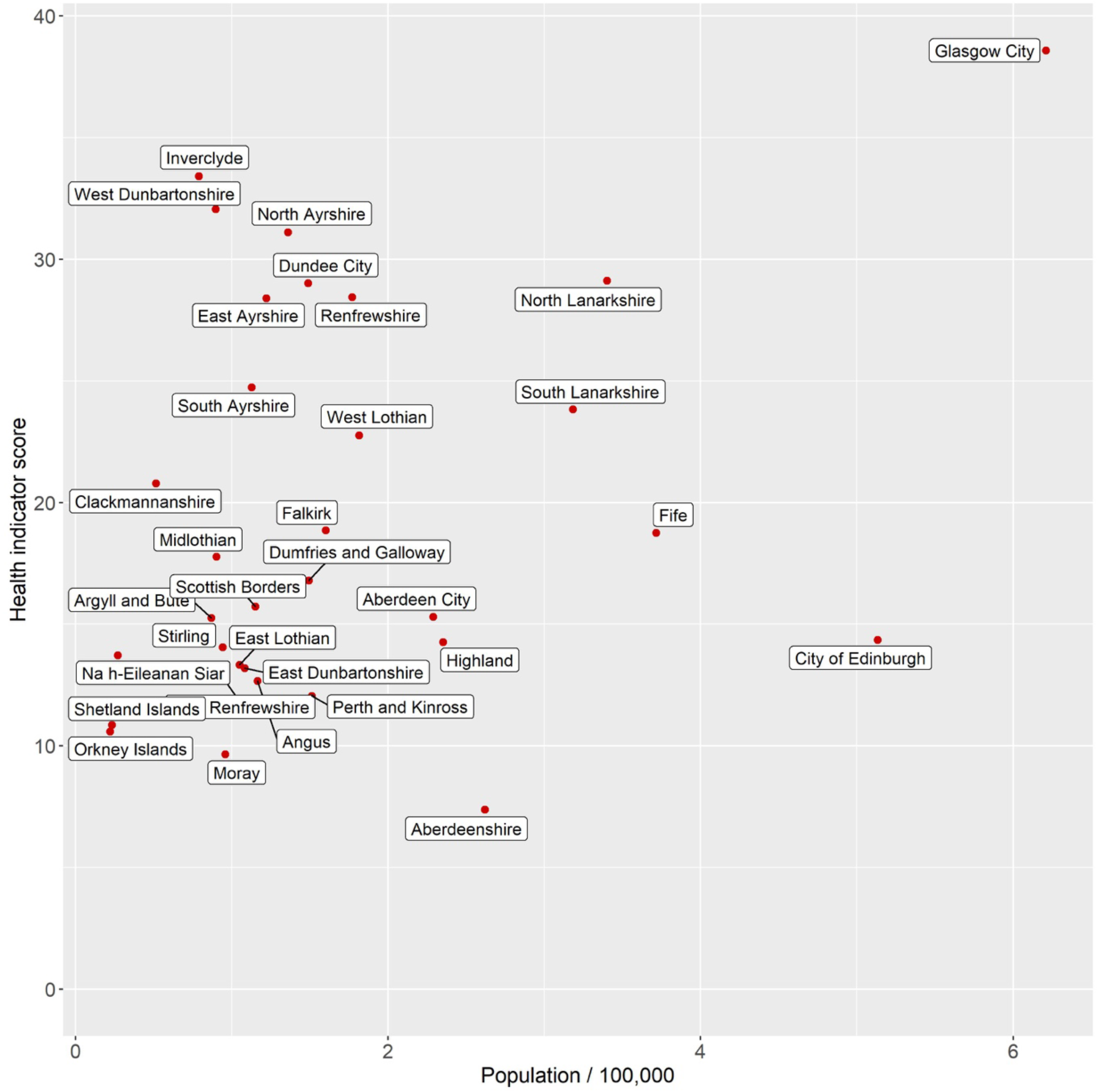
Health indicator against the population for the Scottish Couuncil areas.

### Conclusions

Based on Covid-19 associated deaths from outside care homes, we conclude that whilst deprivation overall (as measured by the Scottish Index of Multiple Deprivation) is significantly associated with increased Covid-19 mortality we have disaggregated this further:

1. Population level risk of Covid-19 mortality is associated with the SIMD indicator that describes (good) accessibility and orthogonally with the SIMD indicator that describes (poor) health. Indicating that areas with poorest health and good access experienced higher Covid-19 mortality.

Risk of excess Covid-19 mortality (Covid-19 deaths as a fraction of all deaths) is most closely associated with the access indicator component of SIMD. This indicates that the areas that have good local connectivity and transport will have higher rates of Covid-19 transmission.

## Appendix II. Model Equations

For individuals in each OA labelled “*i*”, the frequency dependent force of infection *Λ*_*i*_(*t*) is given by: Where:

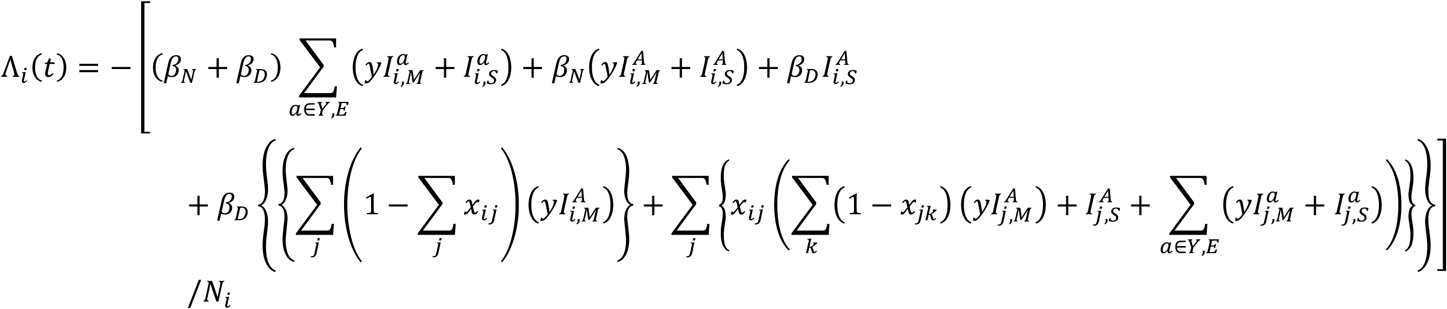

With

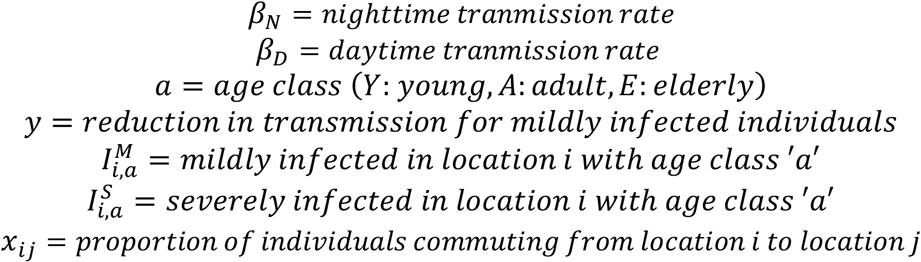

In the compartmental model are infection classes *S* (susceptible), *E* (exposed), *I*^*M*^ (mildly infected), *I*^*S*^ (severely infected), *H* (hospitalised). Model equations for individuals residing in one OA (labelled *i*) and for age class *a’* are therefore:

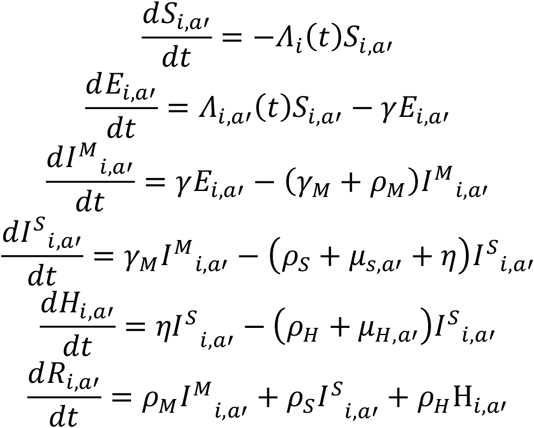

With the number of deaths determined by

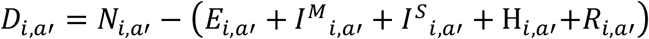

The transition rates are given by:

- γ for *E* to *I*^*M*^,
- ρ_M_ for *I*^*M*^ to *R*,
- ρ_S_ for *I*^*S*^ to *R*,
- ρ_H_ for *H* to *R*
- *γ*_*M*_ for *I*^*M*^ to *I*^*S*^,
- ρ_S_ for *I*^*S*^ to *R*
- η for *I*^*S*^ to *H*,

and the age-dependent mortality rates are given by the values of μ.

### NB

In order to improve the efficiency of the inference, movements of commuters between OAs were batched into groups of 5, with movements between OAs of fewer than five individuals per day, retained at a proportionate rate (i.e. 0.80 of all movements between OAs involving 4 individuals were retained, with the remaining discarded at random). While this reduces the overall network link density, the effect on transmission dynamics is expected to be small. We note that this means that interpretation of the combined *β*_*N*_ and *β*_*D*_ must be made with caution.

## Appendix III. Simulation details

Each simulation takes approximately 2-3 minutes to run. The inference framework is run on a distributed application framework (Akka)^22^. Running Akka on a Cloud Computing IAAS infrastructure (Amazon AWS^23^) allows for rapid scaling upwards to 16Gb and 4 cores per computer node and outwards to 200 computer nodes. In the inference framework each “generation” of the ABC-SMC therefore is complete in approximately 10-20 minutes, with the tolerance in the acceptance scheme usually stabilising after no more than 20 generations (indicating a ‘settled’ posterior estimate). For the inference reported here, this means that the ABC-SMC scheme requires 5×10^4^ - 10^5^ model simulation runs before model convergence (i.e. model tolerance ceases to change substantially between SMC generations.

The model code has been written using industry grade software engineering practices, including Agile development for project task planning^24^, Test Driven Development (TDD)^25^, Pair programming and code reviews to produce unit tested, robust and reusable software components. The majority of the code has been reviewed by a second software developer, with review of the remainder ongoing.

Relevant software developed for this project are deposited here: https://github.com/Kao-Group/SCoVMod

## Appendix IV. Generate of commuter movement patterns

From the current population estimates we draw the number of individuals whose primary residence is mapped onto OA^26^, with their age group. The smallest geographic unit provided publicly by NRS is the Intermediate Zone (IZ)^27^, of which there are approx. 1,200 units in Scotland each with a population of 2,500–6,000 household residents. We refine this, by synthetically distributing individuals down to Census OAs, of which there are approx. 46,000, each with a household population of 100–500. The total population of Scotland from this Census is 5438054 (Young, 919,580; Adult 3,492,421; Elderly 1,026,053). Of the adults, 1,960,712 commute to work (reduced to 647, 034 under lockdown (see details below).

The data for assignment of individuals to work locations is drawn from the NRS Census Flows data^28^, Table WU01UK, which provides origin/destination workplace data for the population from the 2011 census. This is also provided at IZ level, which we distribute to OA level.^29^

The data for adjusting daily movements for the period after lockdown is taken from Google’s Community Mobility Reports^30^, which provide estimates of the proportionate decrease in mobility against a pre-lockdown baseline.

The population and census data were retrieved on 1^st^ April 2020, and the mobility data is updated every few days (last update 1^st^ May 2020).

### Commuter patterns

To model commuter patterns between OAs we construct a commuter network consistent with commuter patterns and density of working age adults recorded in the Scotland census.

Sources from the 2011 Scotland census^31^ were combined to create a semi-synthetic network of work-related movements between households and workplaces. The census provides the number of individuals commuting between ‘place of residence’ and ‘place of work’ at a larger scale than the OA patches used in our model (Table ID WU01SC_IZ2011_Scotland). To generate commutes between the smaller ‘OA’ scale we assigned them a residence OA from within their ‘place of residence’ selected randomly with probability proportional to its working-age household population (Table ID LC1109SC), and a workplace OA from within their ‘Place of work’ selected randomly with probability proportional to the workforce population (Table ID WP101SCoa).

### Movement simulation

The purpose of the movement simulator (“moveSim”) is to provide population and movement input to SCoVMod. Input is in the form of a national population, drawn from census data, and a set of movements—where each individual travels in each time period—drawn from census flows data and modified by mobility data. We assume in this model that the pattern of commuting captures the most important features of. Also, after lockdown, this effect will be enhanced; with evidence from other countries of non-work activity occurring at both reduced volumes [REF google] and at a reduced spatial distance.^32^

### Population

The first output of moveSim is a national population of individuals, each with a unique ID, a starting location, and an age group. At this point we also decide if each individual goes to work and where this occurs, assign them a work location.

### Network Algorithm

The algorithm for assignment of individuals to home locations and age categories applies a unique ID for each individual in the national population. Home locations and age groups are assigned proportionately to the estimated OA population size.

An individual’s workplace is assigned by distributing a proportion of the population of each location to each work location, weighted by the proportion of individuals from each home location in the census flows data who work in another location. For each origin *o* and destination *d* we assign a weight *w*_*o,d*_ from the census flow data: 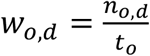 where *n*_*o,d*_ is the total number of people who move from *o* to *d* to work, and *t*_0_ is the total number who move from origin *o* to any location for work.

We take the individuals of each home location if they are eligible to work (total *n*_*o*_); in this case we assume all individuals of adult age 16–65. Each destination is assigned to *n*_*o*_ × *w*_*o,d*_ of these individuals. The individuals who remain have no assigned workplace—either they do not work, or they work within their home location.

### Output format

The population generator of moveSim outputs two tables: the first with *PersonID, Origin, Age* as input to SCoVMod, and the second *PersonID, Origin, Age, Worker, Destination* as input to the moveSim movement simulator. *PersonID* is a unique integer identifier, *Origin* and *Destination* are string location identifiers, *Age* is a categorical identifier in [*Young, Adult, Elderly*], and *Worker* a Boolean identifier indicating an eligible worker.

### Movement

The movement simulator produces input for SCoVMod, in the form of the set of individuals who move from each location in each time step of the simulation. In this case, we use two time steps per day. In the first, workers move to work, and in the second they return home. The simulation is generated stochastically, with a Poisson distributed number of workers moving from each origin to each destination per day, distributed according to the census flows and weighted by population as described above. The volume of movement is reduced uniformly across the population according to the proportional decrease provided by the mobility data.

We also introduce an optimisation to reduce the number of movements that need to be handled by SCoVMod. The number of movements are trimmed to one in five and therefore daytime transmission rates are correspondingly assumed to be per 5 infected individuals. In simulation, there will be many OAs where fewer than five infected individuals would move to them in a given time step, and thus this process of movement thinning would result in many locations not being exposed to infection that would have been with the full movement pattern. Given the low level of infection over the considered scenarios, we assume that the trade-off between increased transmission rate per movement, and reduced movements, will have negligible impact on outcomes.

### Movement Algorithm

For each day of the simulation we consider two time steps: a *day* step where individuals can move to their place of work, and a *night* step where those individuals move back to their home location.

In each day step, we take each destination location *d*. Let *λ*_*d*_ be the number of eligible workers who may move to the destination location. For each day the sampled number who move *s* is drawn from a Poisson distribution:

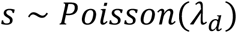

The sampled number of moves *s* is then scaled according to the per cent change in mobility *m*for the given day:

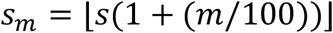

The number of moves is then trimmed 4 in 5 by drawing from a Binomial distribution:

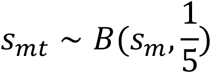

If the sampled number of workers *s*_*mt*_ is less than or equal to the number of workers who may normally move to destination *d*, then those who move are sampled randomly from those who may normally move. However, if *s*_*mt*_ is greater than the number of workers who may normally move to *d*, then the additional workers are drawn randomly from workers who have no assigned destination location. The sampled PersonIDs for each destination location are then collected for output, for each day. For each night of the simulation, the workers who moved in the day step are moved back to their origin location.

### Output format

Each time step of the simulation is output in JSON format for input to SCoVMod. Each time step contains the set of destination location IDs, each containing the set of PersonIDs who move to them.

## Appendix V. Evaluation of validity of google mobility data in rural areas

Reduction in mobility is estimated based on data provided by Google.^33^ In the documentation, it is stated that “Location accuracy and the understanding of categorized places varies from region to region, so we don’t recommend using this data to compare changes between countries, or between regions with different characteristics (e.g. rural versus urban areas).”^34^

In order to provide confidence that inclusion of reductions across regions is appropriate, we here assume that urban areas such as Glasgow and Edinburgh are likely to be well represented, but that rural areas may be less so. To check this, we compare an independent dataset on independent sailings and passenger numbers for ferry services run by Caledonian MacBrayne, who operate all ferry services in the west of Scotland. A comparison of data from 2019 to 2020 and to Google Mobility data, shows a strong fidelity between the two datasets, as well as a substantial reduction in activity at point of lockdown. The similarity prior to lockdown between 2019 and 2020 also suggests that patterns of increased summer activity are unlikely to have had strong influences on our assumptions regarding commuter movements, at least in this area.

**Figure A.V.i.**
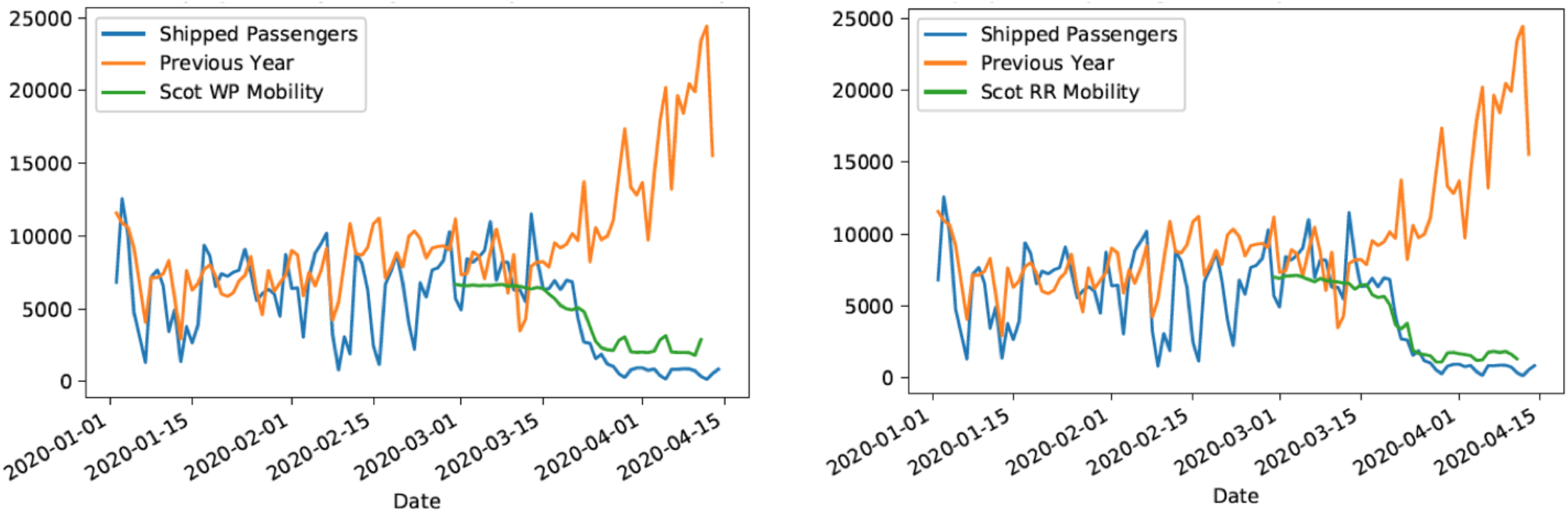
Comparison of Google mobility data for Scotland to CalMac Ferry records. (L) Comparison to workplace mobility (R) Comparison to Recreation mobility. The comparison is relative to the mean value prior to lockdown on March 23rd, 2020.

## Appendix VI. Parameter Posteriors

**Figure A.VI.1.**
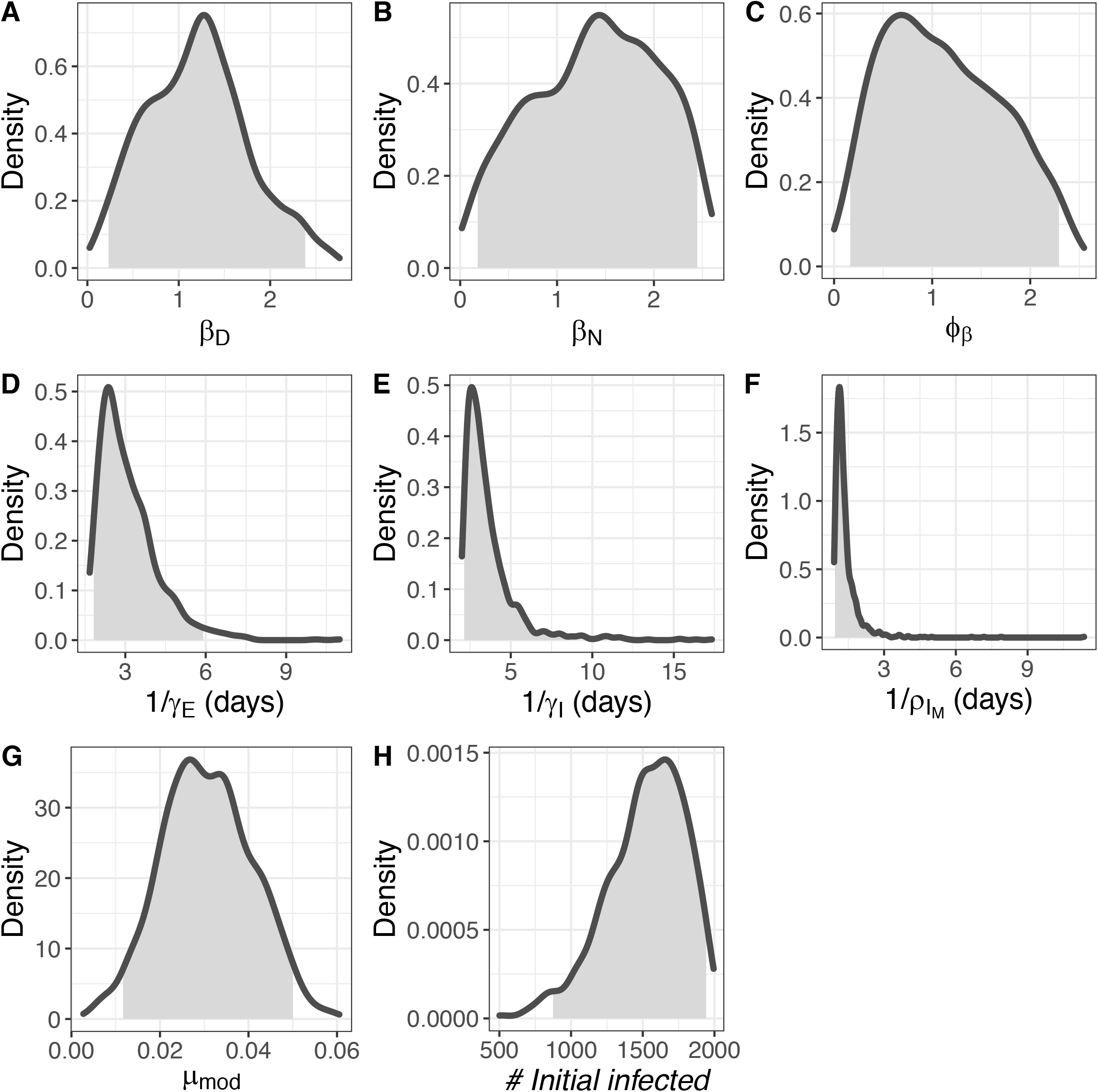
Posterior distributions of fitted parameters. From top L to lower R: (A) frequency dependent transmission rate for severely infectious individuals in “day” locations (per 5 severely infectious individuals, per half day), (B) frequency dependent transmission rate for severely infectious individuals in “night” locations (per severely infectious, per half day), (C) multiplier for mildly infectious individuals). (D) duration of the exposed stage of infection (in days), (E) duration of the mildly infectious period in the absence of recovery (in days), modifier (F) time to recovery for mildly infectious individuals in the absence of progression to severely^i^ infected (days) (G) mortality rate modifier, and (H) number of seed infections. Shaded areas show 95% credible intervals for all parameters. Shaded areas represent 95% credible intervals for the posterior estimates.

**Figure A.VI.2.**
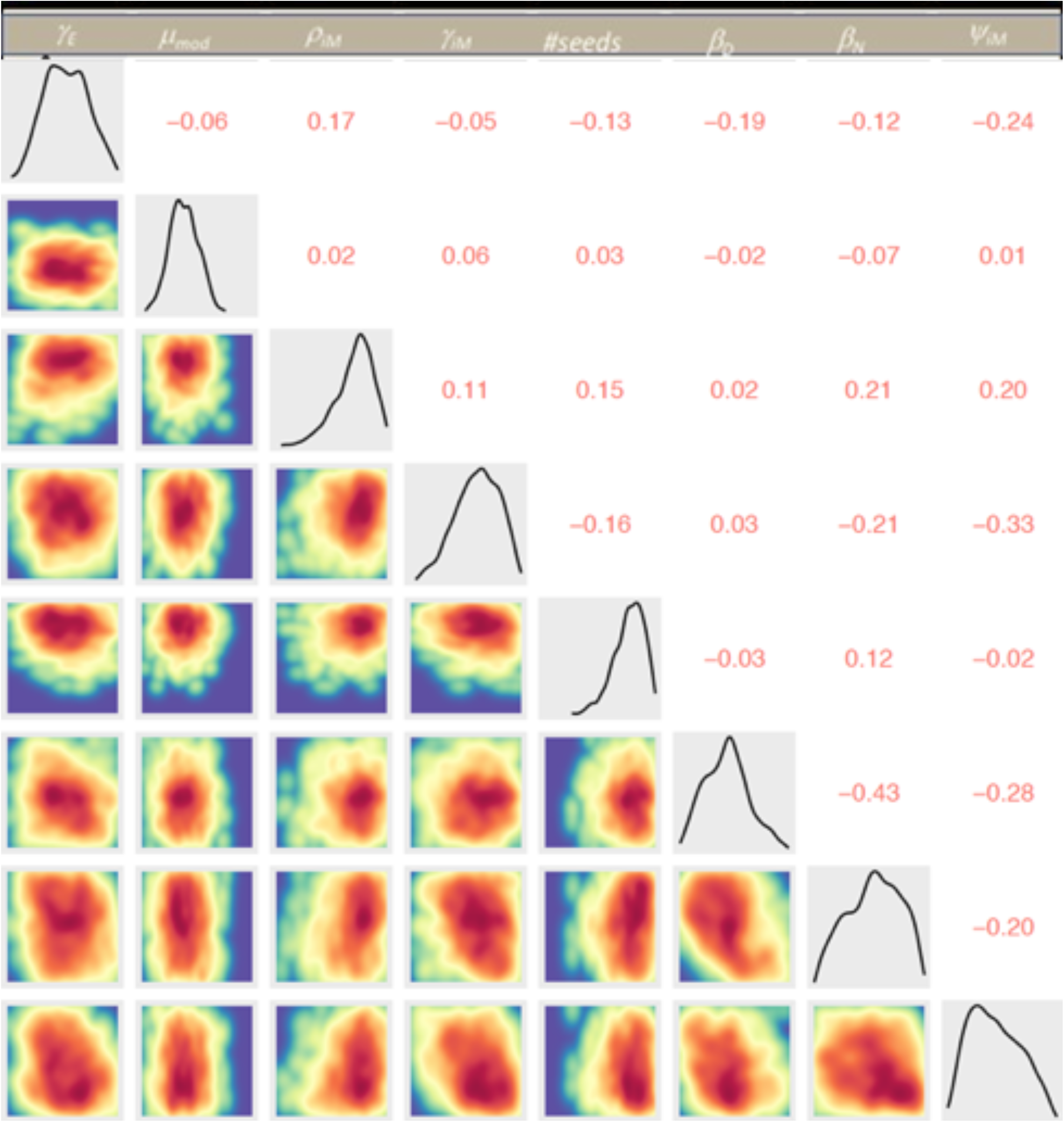
Correlation plots for all parameters.

## Appendix VII. BBC Disclosure, 11^th^ May 2020. Effect of early lockdown & BBC Reporting Scotland, 30^th^ June, 2020

An earlier version of this model which did not include consideration of health index, was used in the BBC One “Disclosure” programme aired at 8:30 pm on Monday, 11^th^ May 2020. While the model results from the refined version are slightly different, the overall conclusions, and estimated number of deaths averted are highly consistent. Using the earlier fitted model, we also explore the impact of an early lockdown, starting at the beginning of the simulation period on March 8^th^, two weeks prior to the actual lockdown rate. In this version, if we assume that the reproduction rate declines below one from two weeks after the imposition of lockdown (the same assumption as for the baseline scenario), we predict on average 577 deaths (95% of simulations within 343 to 1141 deaths) by 26^th^ April 2020, compared to 3259 (range 1736 to 6983) in the baseline scenario (observed number is 2,795, assuming that all deaths occur in the week prior to the week the death is registered in).

One 30^th^ June, a follow-up report on “Reporting Scotland” included an estimate of the impact of a “Swedish-style” lockdown. A recent paper (Flaxman, S., Mishra, S., Gandy, A. et al. Estimating the effects of non-pharmaceutical interventions on COVID-19 in Europe. Nature (2020). https://doi.org/10.1038/s41586-020-2405-7) provides estimates of the reproduction number of COVID-19 in Sweden, before and after introduction of restrictive measures to COVID-19. We assumed an equivalent proportionate reduction in Scotland for a ‘Sweden-like’ control scenario, reducing all transmission rates by the same … amount at the same point (6^th^ April) when we assume Scotland’s more comprehensive lockdown occurred. For an estimate for *R*^*t*^ in Sweden at 2.8 before restriction it is reduced to 0.76 after restrictions. i.e. a factor of 0.27. We conduct 50 simulations (drawing a different parameter set from the fitted posterior for each simulation). For the two scenarios (lockdown as fitted or the baseline scenario, vs. reduction as in the Swedish model). For the ‘baseline’ scenario, this results in an estimated 4523 deaths to the week of June 29^th^ averaged over 50 simulations. If we adopted the Swedish scenario, the simulations suggest that, for the given reduction of 0.27, approximately half of all simulations represent a post-restriction R value > 1, with an average of 39613 deaths (this was in 28 of 50 simulations, or 56%) while in the remainder R < 1, with a mean of 7280 deaths (22 of 50, or 44% of the simulations).

**Figure A.VII.i.**
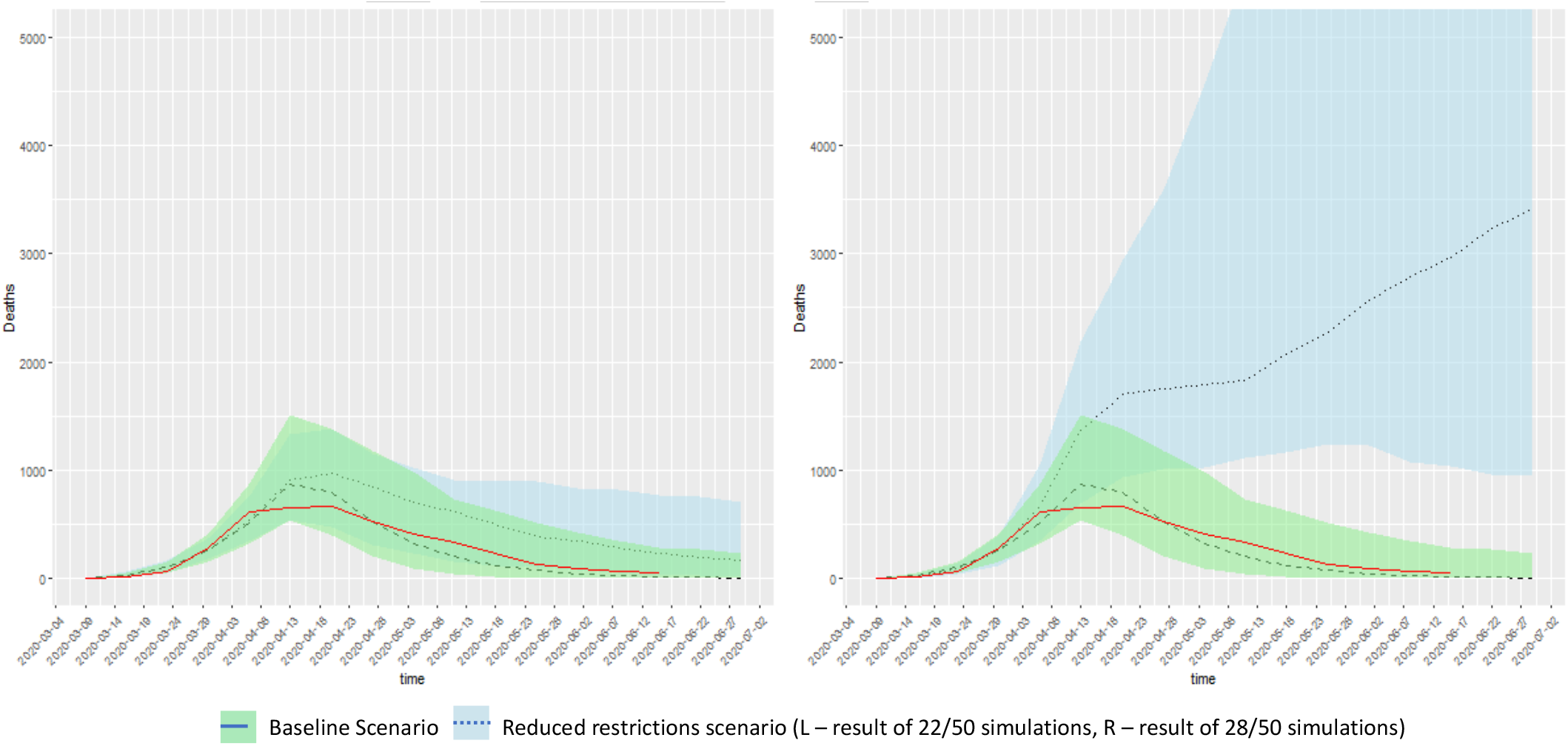
Comparison of ‘baseline scenario’ of Scotland COVID-19 death incidence to a reduced restriction scenario consistent with Swedish controls. On the left, comparison to 22 of 50 simulations with fewer than 14,000 deaths; on the right, comparison to the remaining 28 simulations where deaths exceeded 14,000. In red are the observed data to 17^th^ June, 2020.

For comparison, we also show the ‘early lockdown’ scenario of figure 7 in the same format (Fig. A.VII.ii)

**Figure A.VII.2.**
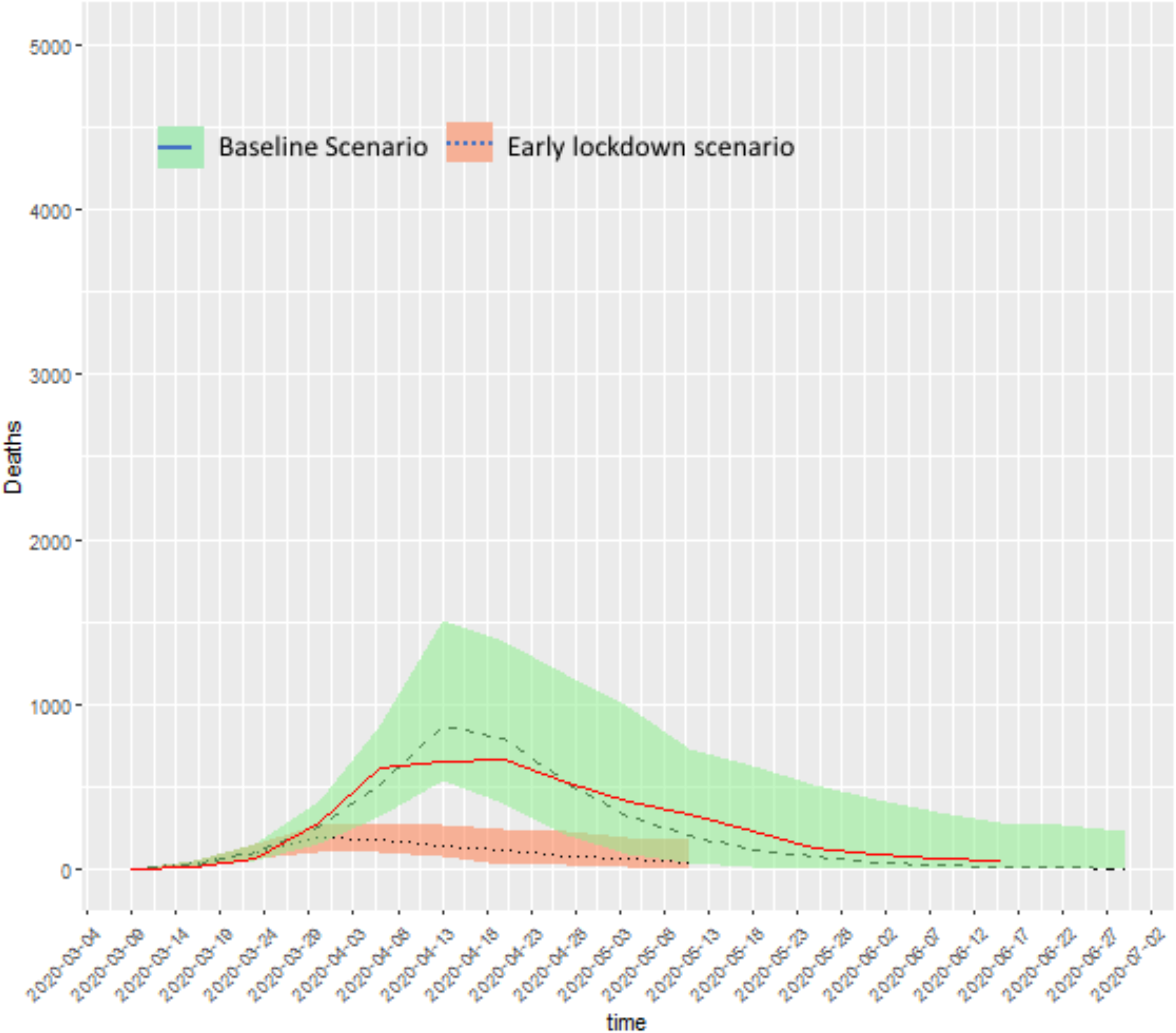
Early lockdown scenario compared to baseline, with true incidence shown in red.

## Appendix VIII. Author contributions

**Banks C.J.*, Colman E. *, Doherty T. *, Tearne O. *, Balaz, D., Arnold M., Atkins, K., Beaunée, G., Bessell P., Enright J., Kleczkowski A, Rossi, G., Ruget A.-S., Kao R.R.†**

CB and EC generated synthetic movement patterns, TD and OT developed and unit tested the code, and developed the ABC-SMC inference scheme. CB provided the Calmac Ferry data comparison. MA provided oversight at APHA. GB advised on ABC-SMC inference and conducted the literature review, PB conducted the statistical analysis of SIMD data, JE contributed to generation of synthetic movement patterns and provided input on model structure throughout the project. DB provided code review. AK and KA provided advice on model development. GR proposed parameter point estimates and prior distributions, ASR generated plots and conducted statistical analyses, RRK conceived the project, and supervised the overall project. CB, EC, TD, and RRK wrote the initial manuscript draft. All authors have read and critiqued the manuscript. RRK is the author for correspondence.

## Footnotes

8 https://www.gov.scot/news/coronavirus-covid-19/

9 https://www.gov.scot/publications/coronavirus-covid-19-framework-decision-making-scotlands-route-map-through-out-crisis/

10 https://www.isdscotland.org/Products-and-Services/EDRIS/COVID-19/index.asp

11 NRS statistics: https://statistics.gov.scot/

12 https://www.scotlandscensus.gov.uk/variables-classification/output-area-2011

13 https://www.nrscotland.gov.uk/files//geography/2011-census/geography-bckground-info-comparison-of-thresholds.pdf

14 https://www.gov.scot/collections/scottish-index-of-multiple-deprivation-2020/

15 https://www.gov.scot/publications/scottish-neighbourhood-statistics-data-zones-background-information/pages/5/#3

16 Google Community Mobility Reports: https://www.google.com/covid19/mobility/

17 Google Community Mobility Reports: https://www.google.com/covid19/mobility/

18 Covid-19: Framework for Decision Making, Further Information (published 23^rd^ April, 2020), accessed 11^th^ May, 2020 https://www.gov.scot/publications/coronavirus-covid-19-framework-decision-making-further-information/

19 Data subject to data sharing agreement due to potential for individual disclosure; it is publicly available at a less resolved level spatially https://www.nrscotland.gov.uk/statistics-and-data/statistics/statistics-by-theme/vital-events/general-publications/weekly-and-monthly-data-on-births-and-deaths/deaths-involving-coronavirus-covid-19-in-scotland

20 https://www.scot.nhs.uk/organisations/

21 http://ide.mit.edu/sites/default/files/publications/Interdependence_COVID_522.pdf

22 https://akka.io

23 https://aws.amazon.com

24 Kent Beck; James Grenning; Robert C. Martin; Mike Beedle; Jim Highsmith; Steve Mellor; Arie van Bennekum; Andrew Hunt; Ken Schwaber; Alistair Cockburn; Ron Jeffries; Jeff Sutherland; Ward Cunningham; Jon Kern; Dave Thomas; Martin Fowler; Brian Marick (2001). “Manifesto for Agile Software Development”.

25 Beck, K., Test-Driven Development By Example, Addison-Wesley, Boston, MA, USA, 2003.

26 Geographic units defined by the ONS: https://www.ons.gov.uk/methodology/geography/ukgeographies/censusgeography

27 NRS definition of geographic units:https://statistics.gov.scot/atlas/resource?uri=http://statistics.gov.scot/id/statistical-geography/S92000003

28 Office for National Statistics, 2011 Census: Special Workplace Statistics (United Kingdom) [computer file]. UK Data Service Census Support. Downloaded from: https://wicid.ukdataservice.ac.uk

29 https://www.scotlandscensus.gov.uk/variables-classification/sns-data-zone-2011

30 Google Community Mobility Reports: https://www.google.com/covid19/mobility/

31 Census data: https://www.scotlandscensus.gov.uk/ods-web/data-warehouse.htmla

32 https://www.covid-19-mobility.org

33 https://www.google.com/covid19/mobility/index.html?hl=en

34 https://www.google.com/covid19/mobility/data_documentation.html?hl=en

